# Breast cancer risks associated with missense variants in breast cancer susceptibility genes

**DOI:** 10.1101/2021.09.02.21262369

**Authors:** Leila Dorling, Sara Carvalho, Jamie Allen, Michael T. Parsons, Cristina Fortuno, Anna González-Neira, Stephan M. Heijl, Muriel A. Adank, Thomas U. Ahearn, Irene L. Andrulis, Päivi Auvinen, Heiko Becher, Matthias W. Beckmann, Sabine Behrens, Marina Bermisheva, Natalia V. Bogdanova, Stig E. Bojesen, Manjeet K. Bolla, Michael Bremer, Ignacio Briceno, Nicola J. Camp, Archie Campbell, Jose E. Castelao, Jenny Chang-Claude, Stephen J. Chanock, Georgia Chenevix-Trench, NBCS Collaborators, J. Margriet Collée, Kamila Czene, Joe Dennis, Thilo Dörk, Mikael Eriksson, D. Gareth Evans, Peter A. Fasching, Jonine Figueroa, Henrik Flyger, Marike Gabrielson, Manuela Gago-Dominguez, Montserrat García-Closas, Graham G. Giles, Gord Glendon, Pascal Guénel, Melanie Gündert, Andreas Hadjisavvas, Eric Hahnen, Per Hall, Ute Hamann, Elaine F. Harkness, Mikael Hartman, Frans B.L. Hogervorst, Antoinette Hollestelle, Reiner Hoppe, Anthony Howell, kConFab Investigators, SGBCC Investigators, Anna Jakubowska, Audrey Jung, Elza Khusnutdinova, Sung-Won Kim, Yon-Dschun Ko, Vessela N. Kristensen, Inge M.M. Lakeman, Jingmei Li, Annika Lindblom, Maria A. Loizidou, Artitaya Lophatananon, Jan Lubiński, Craig Luccarini, Michael J. Madsen, Arto Mannermaa, Mehdi Manoochehri, Sara Margolin, Dimitrios Mavroudis, Roger L. Milne, Nur Aishah Mohd Taib, Kenneth Muir, Heli Nevanlinna, William G. Newman, Jan C. Oosterwijk, Sue K. Park, Paolo Peterlongo, Paolo Radice, Emmanouil Saloustros, Elinor J. Sawyer, Rita K. Schmutzler, Mitul Shah, Xueling Sim, Melissa C. Southey, Harald Surowy, Maija Suvanto, Ian Tomlinson, Diana Torres, Thérèse Truong, Christi J. van Asperen, Regina Waltes, Qin Wang, Xiaohong R. Yang, Paul D.P. Pharoah, Marjanka K. Schmidt, Javier Benitez, Bas Vroling, Alison M. Dunning, Soo Hwang Teo, Anders Kvist, Miguel de la Hoya, Peter Devilee, Amanda B. Spurdle, Maaike P.G. Vreeswijk, Douglas F. Easton

## Abstract

**BACKGROUND:** Protein truncating variants in *ATM, BRCA1, BRCA2, CHEK2* and *PALB2* are associated with increased breast cancer risk, but risks associated with missense variants in these genes are uncertain.

**METHODS:** Combining 59,639 breast cancer cases and 53,165 controls, we sampled training (80%) and validation (20%) sets to analyze rare missense variants in *ATM* (1,146 training variants), *BRCA1* (644), *BRCA2* (1,425), *CHEK2* (325) and *PALB2* (472). We evaluated breast cancer risks according to five *in-silico* prediction-of-deleteriousness algorithms, functional protein domain, and frequency, using logistic regression models and also mixture models in which a subset of variants was assumed to be risk-associated.

**RESULTS:** The most predictive *in-silico* algorithms were Helix (*BRCA1, BRCA2* and *CHEK2)* and CADD (*ATM*). Increased risks appeared restricted to functional protein domains for *ATM* (FAT and PIK domains) and *BRCA1* (RING and BRCT domains). For *ATM, BRCA1* and *BRCA2*, data were compatible with small subsets (approximately 7%, 2% and 0.6%, respectively) of rare missense variants giving similar risk to those of protein truncating variants in the same gene. For *CHEK2*, data were more consistent with a large fraction (approximately 60%) of rare missense variants giving a lower risk (OR 1.75, 95% CI (1.47-2.08)) than *CHEK2* protein truncating variants. There was little evidence for an association with risk for missense variants in *PALB2*. The best fitting models were well calibrated in the validation set.

**CONCLUSIONS:** These results will inform risk prediction models and the selection of candidate variants for functional assays, and could contribute to the clinical reporting of gene panel testing for breast cancer susceptibility.

## Introduction

Genetic testing for cancer susceptibility is now part of mainstream clinical practice. For breast cancer susceptibility, genetic testing generally focuses on high-risk genes, notably *BRCA1, BRCA2, PALB2* and *TP53*, but testing of larger panels that include so-called “moderate-risk” genes is being increasingly offered (1). While the evidence that many of these genes are risk associated is clear, for most this evidence is based on carrying a protein truncating variant (PTV). Besides PTVs, genetic testing also identifies missense variants for which the impact on protein function and associated cancer risk is generally unknown (“variants of uncertain significance” (VUS)), resulting in a major problem for genetic counselling. Some missense variants have been shown to confer risk (2, 3) with risk estimates comparable to PTVs, and it is possible that missense variants contribute substantially to risk (4, 5), at least in some genes. However, defining the set of missense variants in each gene that may confer risk, and their associated risk estimates, presents an ongoing problem.

Resolving this problem is complex as most variants are individually very rare, so the evidence must be based on combining data across multiple variants in a statistical model. To this end, efforts have been made to develop statistical algorithms that score missense variants according to *in silico* features that may predict pathogenicity. Here, we have compared the usefulness of five *in silico* algorithms in predicting breast cancer risk associated with missense variants using sequenced germline DNA from more than 59,000 cases and 53,000 controls from studies in the Breast Cancer Association Consortium (BCAC) (6) participating in the BRIDGES study (7). We used the most predictive *in silico* algorithm to estimate the risks of breast cancer associated with subsets of rare missense variants, defined by categories of the *in silico* score, in *ATM, BRCA1, BRCA2, CHEK2* and *PALB2*. These predictions were then validated using an independent dataset.

## Methods

### SUBJECTS

We included data from female breast cancer patients (cases) and unaffected controls from 45 studies participating in the BRIDGES collaboration, as previously documented (7). Of these, 30 were population-based or hospital-based studies (hereafter: population studies) including cases and controls sampled independently of family history. A further 14 studies oversampled cases with a family history of breast cancer (hereafter: familial studies). All studies were approved by the relevant ethical review boards and used appropriate consent procedures. Five duplicated samples were identified and removed. After quality control procedures (see below), 53,165 controls and 59,639 cases with an invasive (53,838; 90.3%) or in situ (4,153; 7.0%) tumor, or tumor of unknown invasiveness (1,648; 2.7%), were included in the analyses. Of these, 50,414 controls and 48,230 cases were from population studies.

### LABORATORY METHODS, VARIANT CALLING AND CLASSIFICATION

Of the panel of 34 genes screened in BRIDGES (7), five (*ATM, BRCA1, BRCA2, CHEK2, PALB2*) were chosen for further analysis and presented here. These five genes, where the evidence for association with breast cancer risk is strongest, are most relevant to risk prediction and included in the current version of the BOADICEA/CanRisk risk prediction tool (8). Details of library preparation, sequencing, variant calling, quality control procedures and variant classification have been documented previously (7). Missense variants in the entire gene were identified using the Ensembl Variant Effect Predictor (VEP; version 101.0) (9). Rare variants for *in silico* analysis were defined as those with frequency <0.1% (calculated as previously described (7)); in addition, variants with frequency <0.5% were retained for a frequency-based analysis. Carriers of missense variants predicted to effect RNA splicing, according to the MaxEntScan tool (10) and SpliceAI scores (11), were removed (see Additional File 1: Additional Table 1). Variants were annotated for functional protein domain location, defined according to published literature, the UniProt Knowledgebase (12) and, for BRCA1 and BRCA2, the ENIGMA *BRCA1/2* expert panel guidelines (13) (see Additional File 2: Additional Table 2). Variants were also classified for disease pathogenicity assertion in ClinVar (14) with a filter for no conflicting interpretations; for *BRCA1* and *BRCA2*, variants were also reviewed against the ENIGMA *BRCA1/2* expert panel guidelines. The ENIGMA terminology report (15) reserves use of the word “pathogenic” to describe variants associated with at least a twofold cancer risk; however, for the purpose of this article we describe any variant associated with risk as pathogenic. Variants were scored using five *in silico* prediction algorithms: Align-GVGD (16), Combined Annotation Dependent Depletion (CADD; version 1.4) (17), Rare Exome Variant Ensemble Learner (REVEL) (18), BayesDel (without allele frequency; version 1) (19) and Helix (version 4.2.0) (20). The first four are widely used for variant classification in cancer susceptibility genes. Align-GVGD classifies variants according to the level of cross-species conservation observed for a single missense substitution while considering the biophysical characteristics of the amino acids. CADD, BayesDel and REVEL are ensemble methods that integrate several different annotations, including conservation metrics, regulatory information, transcript information and protein-level scores, into a single score of deleteriousness. Helix combines structural, alignment and gene data with a strict training regime where circularity is actively avoided to produce a variant score and certainty estimate. All variants were scored using default software settings. For Align-GVGD the sequence alignment with the deepest phylogeny level was used. For *BRCA1*, we also annotated variants using the prediction of loss-of-function made by the Saturation Genome Editing (SGE) experiments of Findlay *et al* (21), which involved a comprehensive functional assessment of missense variants lying within the functional domain coding regions of *BRCA1*.

### STATISTICAL ANALYSIS

The dataset was split into a training (80% of individuals) and a validation (20%) set. Samples for the validation set were selected randomly from population studies of cases unselected for family history of breast cancer and controls, in countries contributing a total of >5000 samples (Denmark, Germany, Singapore (Chinese), Sweden, UK, USA). All remaining samples were included in the training set. The training set included 37,211 cases from population studies, 11,409 cases from familial studies and 42,334 controls. Of these, 3,818 individuals were carriers of PTVs in one or more of the five genes under consideration and were excluded from all analyses except the mixture models (see below). The validation set included 11,019 cases and 10,831 controls from population studies and did not include any carriers of PTVs. Oversampling of cases with a family history increases power but leads to biased effect sizes, so we chose this approach to maximize the power to discriminate between models in the training set, which could then be refit and tested on a dataset unselected for family history. All analyses were adjusted for country as a covariate; in addition, for Malaysia and Singapore, the three distinct ethnic groups (Chinese, Indian, Malay) were treated as different strata, and the UK was treated as three strata (SEARCH from East Anglia, GENSCOT from Scotland, and PROCAS and FHRISK from north-west England).

### TRAINING DATASET ANALYSIS

An analysis flow diagram is presented in Additional Figure 1 (see Additional File 2). Analyses were performed in R version 4.0.3 (R: A Language and Environment for Statistical Computing; http://www.r-project.org). We first used logistic regression (LR) to explore which of the five in silico scores (Align-GVGD, BayesDel, CADD, Helix and REVEL - all analyzed as continuous variables) were most strongly associated with risk of breast cancer. For *BRCA1*, we also analyzed the SGE score. These analyses were restricted to carriers of a rare (frequency <0.1%) missense variant in the training set, with an endpoint of breast cancer occurrence (yes/no). The strongest predictors were used to test the association of different categories of the score(s) compared to a baseline category, in conjunction with functional protein domains, and hence create a set of risk categories. LR was then used in the training set (carriers and non-carriers) to estimate the odds ratios (OR) associated with different risk categories. As an alternative approach, we fitted mixture models in which only a proportion of variants (α) was assumed to be risk associated in the given gene; the OR was assumed to be the same for all risk associated variants, but the proportion of risk associated variants varied by risk category (as defined in the LR models). This model is motivated by the binary variant classification approach used in clinical genetics, where all variants are assumed to be either associated with moderate-high risk (likely pathogenic) or not (likely benign) (22). We considered two types of mixture model: a constrained model in which the missense OR was equal to that of PTVs, and an unconstrained model in which the missense OR could differ from the PTV OR. Carriers of PTVs in the gene under consideration were re-included in the mixture models (to allow the risk associated missense OR to be constrained to the PTV OR). The mixture models were fitted using an expectation-maximization (EM) algorithm (23). In the expectation step, the (posterior) probability that each variant was risk associated, given the case control data on that variant in the training set and the current parameter values, was calculated. These probabilities were then used as weights in a logistic regression analysis in the maximization step. In a case-control dataset, the naïve proportions, α, will be biased because risk associated variants are more likely to be found in cases. For the final models, therefore, we also computed the proportions based only on variants reported in controls. To evaluate the overall fit of the models, we compared log-likelihoods.

The initial model selection was based on all samples, but final parameter estimates were obtained from population studies only. In the results, the ORs, *P*-values and α presented are from population studies, unless indicated by the suffix “ALL”.

We evaluated individual risk variants previously reported in literature and, in aggregate, those classified as “pathogenic” or “likely pathogenic” (hereafter, all termed: (likely) pathogenic) according to clinical guidelines. To examine whether variant frequency is associated with risk, we used a case-only LR analysis to test frequency up to 0.5% on a continuous scale and a log scale, and to compare rare variants (frequency < 0.1%) with more common variants (frequency 0.1% - 0.5%). The more common variants were also evaluated individually.

### VALIDATION DATASET ANALYSIS

To evaluate the calibration of the training models, we performed case-control analyses using the validation dataset. In these analyses, OR estimates were fixed according to the population estimates from the training models (Table 1), but the other parameters (intercept and country covariates) were re-estimated, since the case-control proportions might differ between the training and validation datasets. From the validation model, we extracted the predicted probability that each individual was a case and hence derived expected numbers of cases and controls in each risk group. These were used to plot observed versus expected OR estimates and perform a goodness of fit chi-squared test.

**Table 1:**
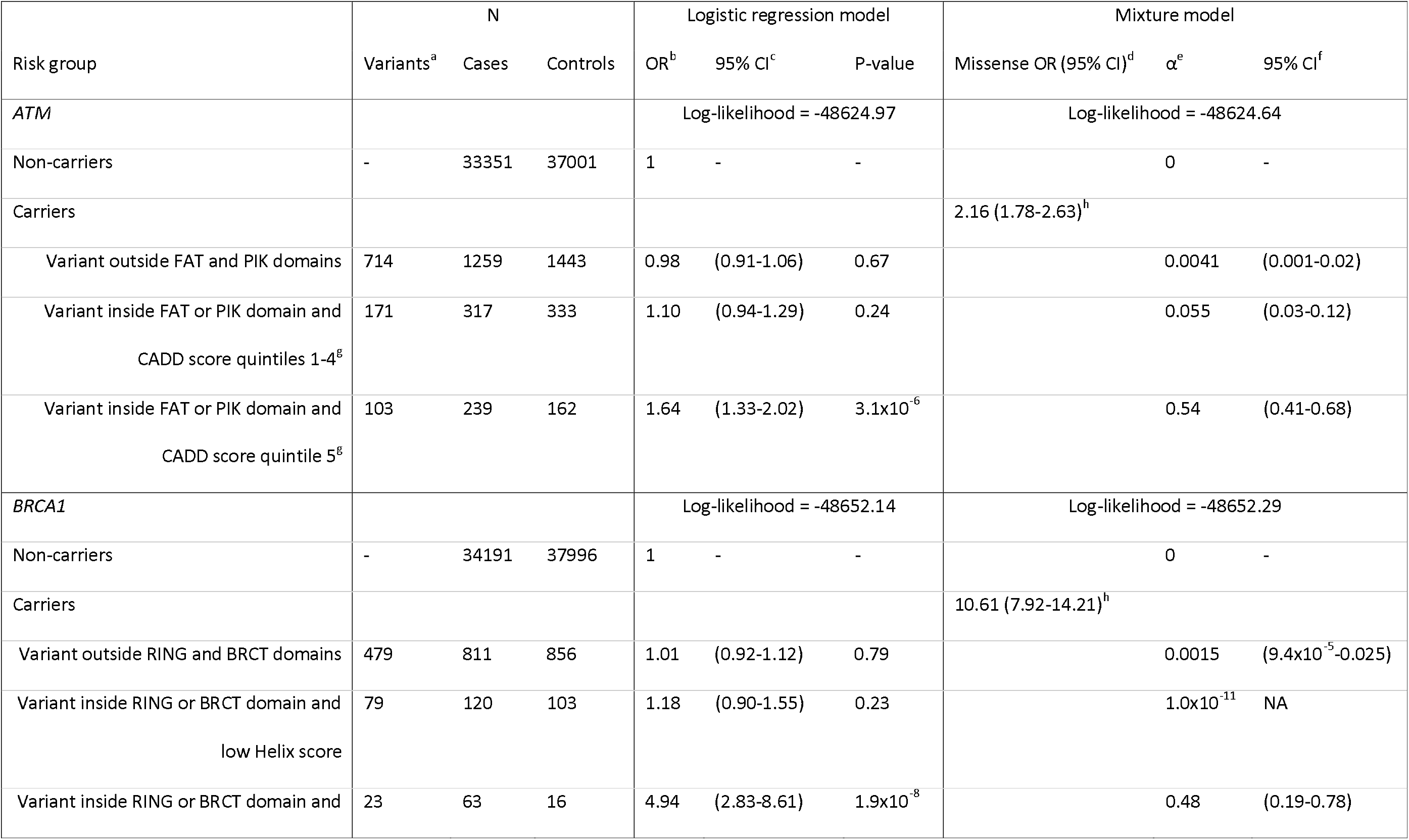
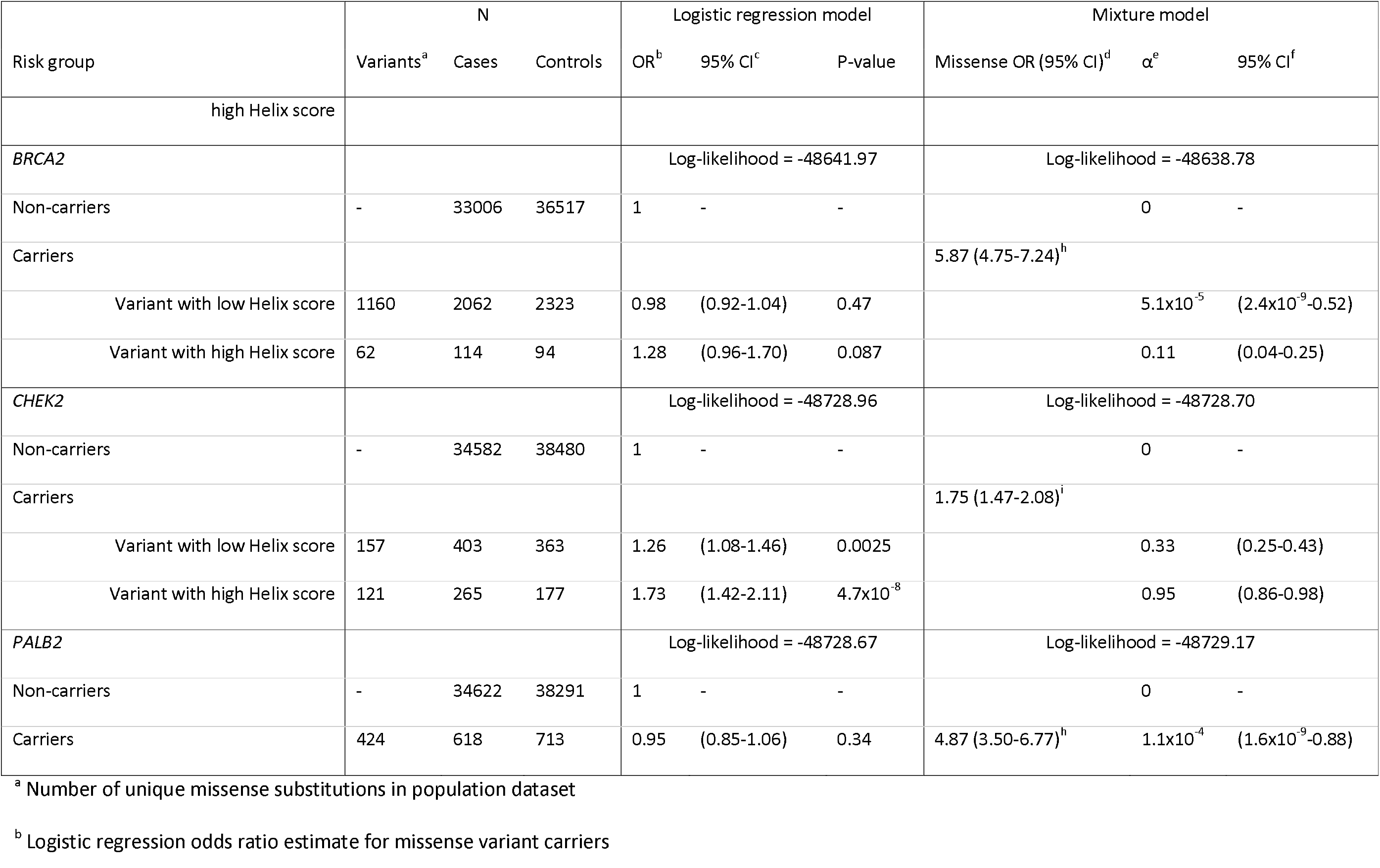
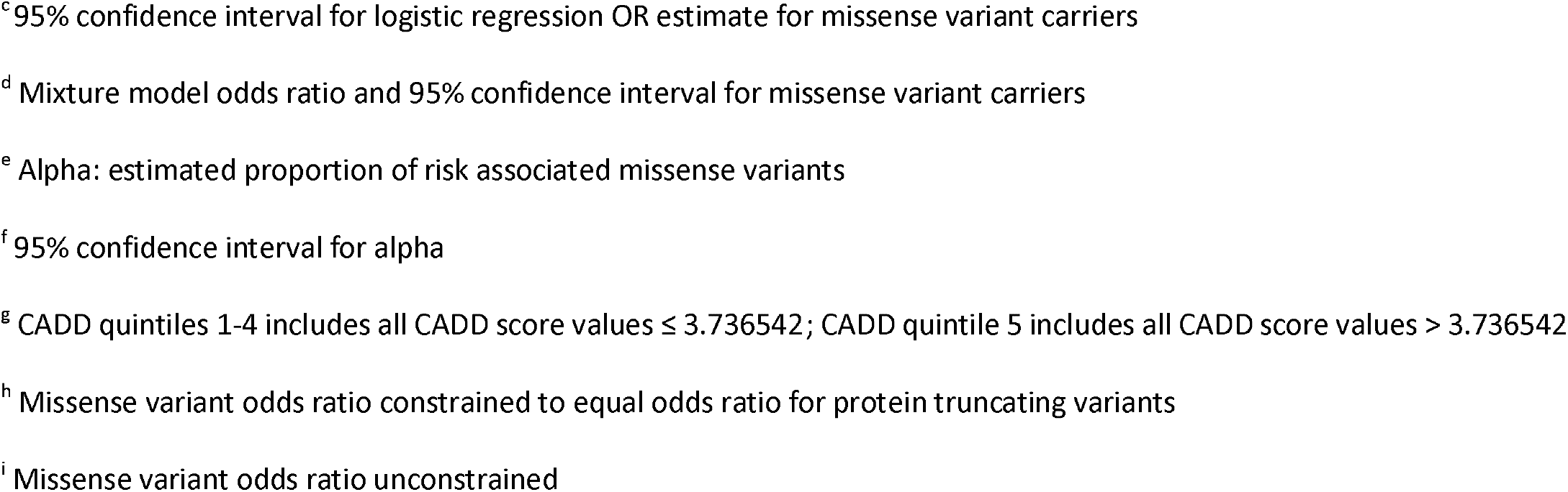
Breast cancer risk association results from logistic regression and mixture models of population training samples

The mixture models were assessed similarly, with the exception that both the OR parameter and the proportion of risk associated variants, α, were fixed. However, an adjustment to α was incorporated to allow for the different distribution of cases and controls within the validation set compared to the training set. To do this, the proportions of cases and controls that were carrying a risk associated variant in the training set were estimated separately and α in the validation set was then computed as a weighted average of these two estimates. As an alternative approach, the predicted ORs in the validation set were computed using the posterior probabilities (PP) of each variant being risk associated (from the training set) as weights. This analysis was restricted to the subset of individuals carrying variants found in the training set or carrying no variant.

As a final analysis, a single unconstrained logistic regression model comprising all the defined risk groups across the five genes, with non-carriers of any missense variant as the baseline group, was fitted, and the risks in the validation set were evaluated.

## Results

### ATM

The analysis of *ATM* missense variants included 4,522 carriers of 1,146 unique variants. In the carrier only analysis, BayesDel (p_ALL_=0.024), CADD (p_ALL_=0.0022), Helix (p_ALL_=0.0045) and REVEL (p_ALL_=0.024) scores were all predictive of risk (see Additional File 1: Additional Table 3). For the most strongly associated score, CADD, the risk appeared to be restricted to the fifth quintile (Q5; CADD > 3.736542; p=0.033 compared with third quintile). Functional protein domain was also predictive, with increased risks associated with the FRAP-ATM-TRRAP (FAT; p_ALL_=9.5×10^−4^) and Phosphatidylinositol 3-kinase and 4-kinase (PIK; p_ALL_=0.0016) domains compared with variants outside a known domain. Including CADD and protein domain, only variants in the category that included CADD Q5 variants in the FAT or PIK domains (FAT/PIK + CADD5) were associated with risk relative to non-carriers (OR 1.64 (1.33-2.02), p=3.1×10^−6^; Table 1, Figure 1a, Figure 2a). In the most parsimonious mixture model, risk associated variants conferred an equivalent risk to PTVs (OR 2.16 (1.78-2.63)); an estimated 54% (95% CI (41%-68%)) of variants in the FAT/PIK + CADD5 risk group were risk associated, compared to less than 6% of variants in other risk categories (Table 1, Figure 1a, Figure 2a). There was no evidence that missense variants were associated with a different risk compared with PTVs (p=0.48). The mixture model was a slightly better fit to the data than the LR model (2 x log-likelihood difference = 0.67).

**Figure 1.**
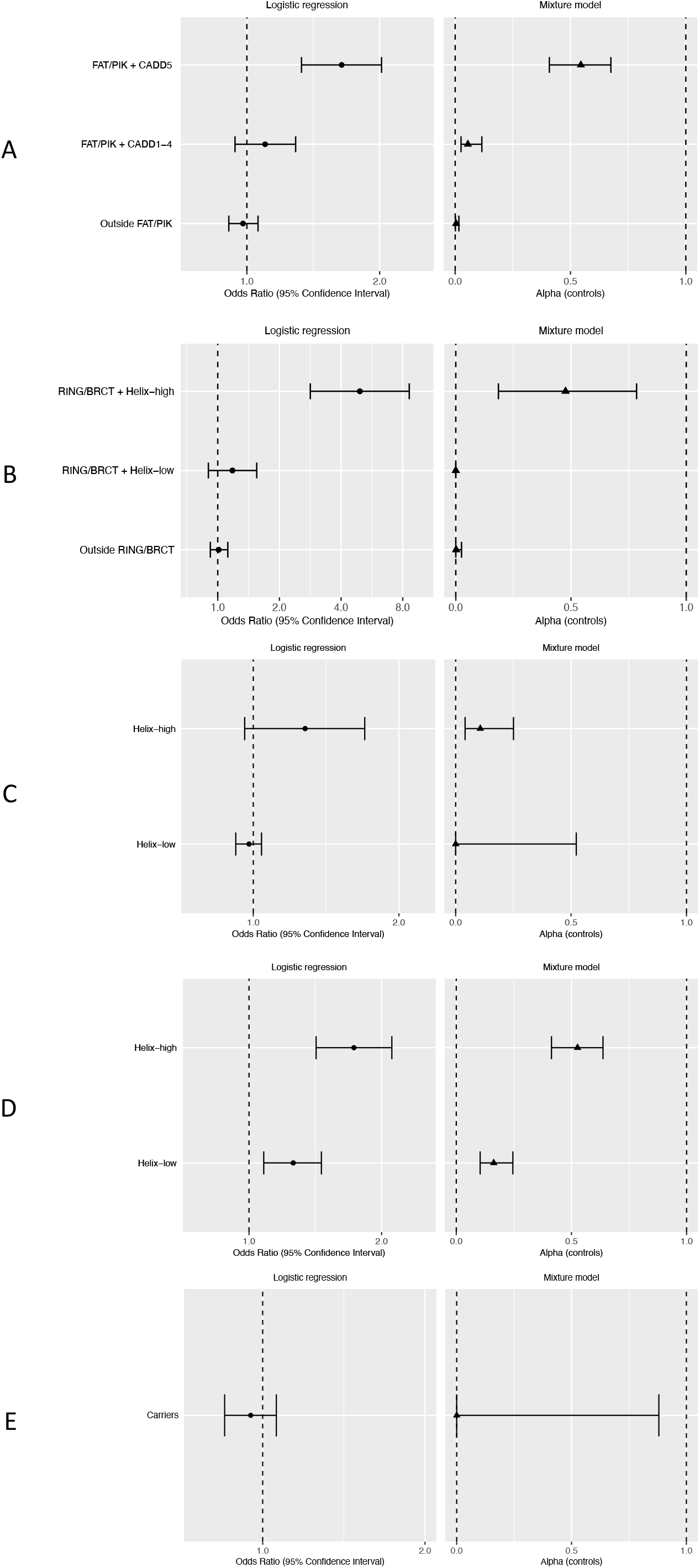
Odds ratios and alpha estimates for each of five genes in population training samples. **Figure 1 panel A legend** ***ATM***. Odds ratios for breast cancer risk from logistic regression models. Alpha is the estimated proportion of risk associated variants from mixture models, based on variants in control samples. *ATM* risk categories: variants lying within the FAT or PI3K/PI4K protein domains with CADD score in the fifth quintile (FAT/PIK + CADD5); variants lying within the FAT or PI3K/PI4K protein domains with CADD score in any of the first four quintiles (FAT/PIK + CADD1-4); variants lying outside the FAT and PI3K/PI4K protein domains (Outside FAT/PIK). **Figure 1 panel B legend** ***BRCA1***. Odds ratios for breast cancer risk from logistic regression models. Alpha is the estimated proportion of risk associated variants from mixture models, based on variants in control samples. *BRCA1* risk categories: variants lying within the RING or BRCT domains with a high Helix score (RING/BRCT + Helix-high); variants lying with the RING or BRCT domains with a low Helix score (RING/BRCT + Helix-low); variants lying outside the RING and BRCT domains (Outside RING/BRCT) **Figure 1 panel C legend** ***BRCA2***. Odds ratios for breast cancer risk from logistic regression models. Alpha is the estimated proportion of risk associated variants from mixture models, based on variants in control samples. *BRCA2* risk categories: variants with a high Helix score (Helix-high); variants with a low Helix score (Helix-low). **Figure 1 panel D legend** ***CHEK2***. Odds ratios for breast cancer risk from logistic regression models. Alpha is the estimated proportion of risk associated variants from mixture models, based on variants in control samples. *CHEK2* risk categories: variants with a high Helix score (Helix-high); variants with a low Helix score (Helix-low). **Figure 1 panel E legend** ***PALB2***. Odds ratios for breast cancer risk from logistic regression models. Alpha is the estimated proportion of risk associated variants from mixture models, based on variants in control samples. *PALB2* risk categories: carriers of any missense variant (Carriers)

**Figure 2.**
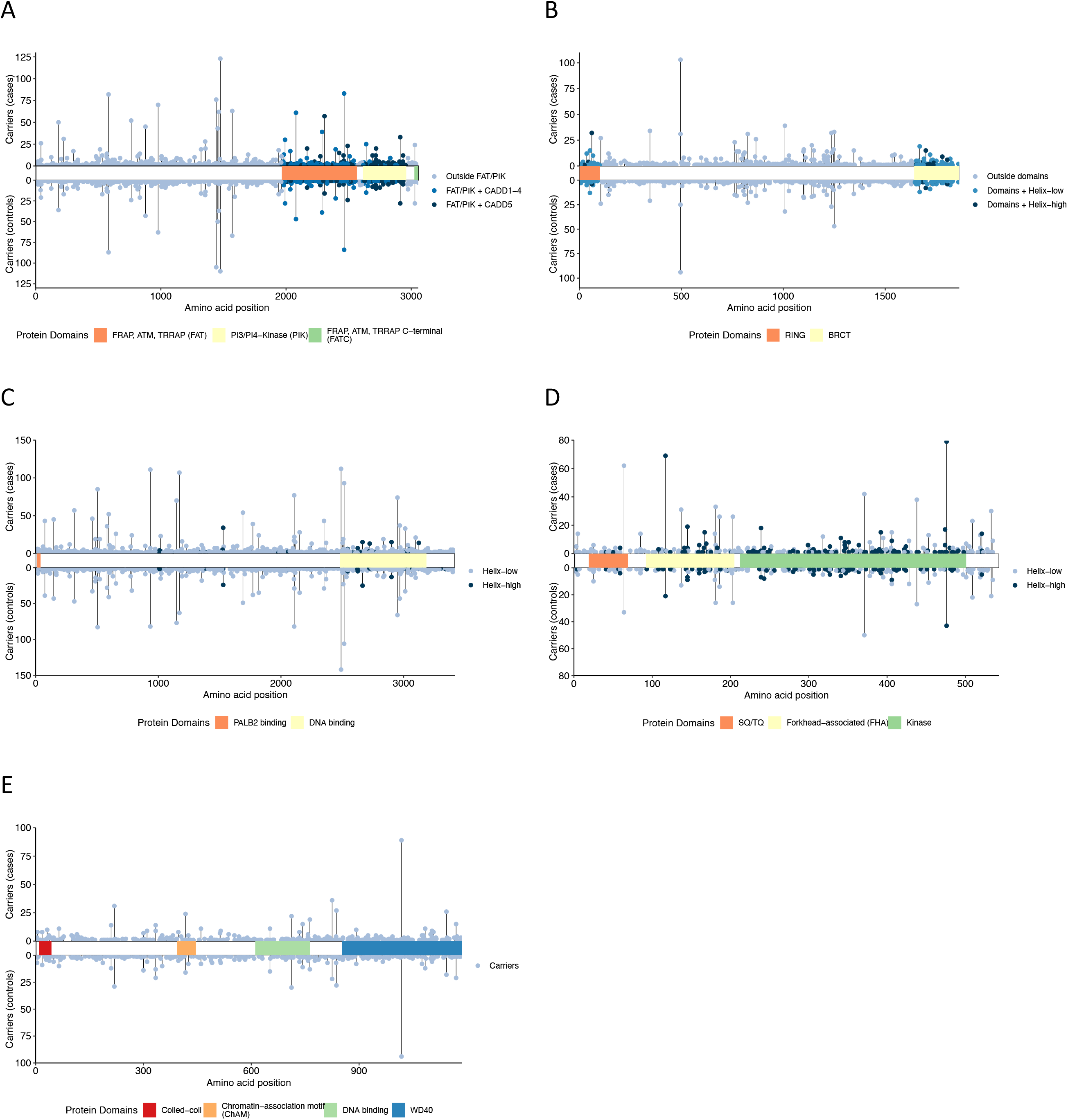
Case and control carriers across all samples for each observed missense variant by gene. **Figure 2 panel A legend** ***ATM***. *ATM* risk categories: variants lying within the FAT or PI3K/PI4K protein domains with CADD score in fifth quintile (FAT/PIK + CADD5); variants lying within the FAT or PI3K/PI4K protein domains with CADD score in any of first four quintiles (FAT/PIK + CADD1-4); variants lying outside the FAT and PI3K/PI4K protein domains (Outside FAT/PIK). **Figure 2 panel B legend** ***BRCA1***. *BRCA1* risk categories: variants lying within the RING or BRCT domains with a high Helix score (RING/BRCT + Helix-high); variants lying with the RING or BRCT domains with a low Helix score (RING/BRCT + Helix-low); variants lying outside the RING and BRCT domains (Outside RING/BRCT). **Figure 2 panel C legend** ***BRCA2***. *BRCA2* risk categories: variants with a high Helix score (Helix-high); variants with a low Helix score (Helix-low). **Figure 2 panel D legend** ***CHEK2***. *CHEK2* risk categories: variants with a high Helix score (Helix-high); variants with a low Helix score (Helix-low). **Figure 2 panel E legend** ***PALB2***. *PALB2* risk categories: carriers of any missense variant (Carriers)

Thirteen *ATM* missense variants were classified as (likely) pathogenic on the ClinVar database (see Additional File 2: Additional Table 4). These variants, in aggregate, were associated with an increased risk (OR 1.85 (0.98-3.50, p=0.060; p_ALL_=0.00053)). However, the association of (likely) pathogenic variants was not present when the analysis was restricted to the five variants not in the FAT or PIK domains (OR=0.97 (0.19-5.08)), though the carrier numbers were small and the confidence interval wide. Conversely, variants in the FAT/PIK + CADD5 risk group, in aggregate, remained risk associated, even when variants defined as (likely) pathogenic were excluded (OR 1.60 (1.29-1.99)). Two of the variants classified as (likely) pathogenic were observed in controls only (Table S4). One of these (c.8546G>C) is located in the PIK domain, the other (c.3848T>C) is not within any domain; however, both have a Q5 CADD score.

The pathogenic variants listed on ClinVar include c.7271T>G (p.Val2424Gly), previously reported as associated with high risk of breast cancer (24, 25). In the training dataset, c.7271T>G was identified in 12 cases (6 population-based) and 6 controls and was not associated with risk (p=0.37, p_ALL_=0.081); its population-based OR estimate of 1.63 (0.56-4.73) was lower than previous estimates (for example (26)). Another variant previously reported as risk associated, c.6919C>T (p.Leu2307Phe) (27), was associated with an increased population risk (OR=3.71 (1.87-7.38), p=0.00018). Both variants are located in the FAT domain and have a CADD score in Q5, but after excluding them from the model there remained a significantly increased risk for carriers in the FAT/PIK + CADD5 risk group (OR 1.48 (1.18-1.85), p=0.00064).

### BRCA1

The analysis of *BRCA1* missense variants included 2,288 carriers of 644 unique variants. For missense variant carriers, all five continuous *in silico* scores were associated with risk (Align-GVGD p_ALL_=1.3×10^−8^, BayesDel p_ALL_=0.0013, CADD p_ALL_=0.011, Helix p_ALL_=2.1×10^−9^, REVEL p_ALL_=1.5×10^−5^). Variants in two protein domains were also significantly associated with risk compared with variants outside these domains (RING finger domain p_ALL_=3.5×10^−4^; BRCA1 C-terminal domains (BRCT I-II) p_ALL_=0.0030; see Additional File 1: Additional Table 3). The Helix tool categorizes variants with a high score (> 0.5) as “deleterious” and variants with a low score (< 0.5) as “benign”; hereafter we refer to these categories as Helix-high and Helix-low, respectively. Including Helix category and protein domain, we found that only variants that were inside the RING or BRCT I-II domains and also in the Helix-high category (RING/BRCT + Helix-high) were associated with risk (OR compared with non-carriers 4.94 (2.83-8.61), p=1.9×10^−8^; p_ALL_=2.5×10^−9^; Table 1, Figure 1b, Figure 2b). In a mixture model in which the OR for risk associated missense variants was constrained to that for PTVs (OR 10.61 (7.92-14.21)), the estimated proportions of risk associated variants in the RING/BRCT + Helix-high risk category was 48% (19%-78%) and close to 0% for all other variants (Table 1, Figure 1b, Figure 2b). There was no evidence that the risk associated missense OR differed from the PTV OR (p=0.98). The LR and mixture models were similarly good fits to the data (2 x log-likelihood difference = 0.30).

According to the ENIGMA guidelines and/or ClinVar classifications, 13 of the *BRCA1* missense variants in the dataset (four in the RING domain and nine in the BRCT domains) would be classified as (likely) pathogenic (see Additional File 2: Additional Table 4). In total, the 13 variants were carried by 60 cases and 6 controls and were strongly associated with risk in the subset of population samples (OR 16.68 (5.16-53.94), p=2.6×10^−6^). In our dataset, the most frequent of these variants was c.181T>G (p.Cys61Gly), carried by 29 cases and 2 controls (OR 15.06 (3.58-63.36)). After excluding all (likely) pathogenic variants, there also remained an increased risk associated with variants in the RING/BRCT + Helix-high category (OR 2.39 (1.19-4.78), p=0.014)).

*BRCA1* Saturation Genome Editing (SGE) score was available for 100 unique variants and was strongly associated with risk (p_ALL_=1.5×10^−4^; see Additional File 1: Additional Table 3). Carriers of variants with an SGE loss of function (LOF_SGE_) consequence had a higher risk than carriers of variants with a functional (FUNC_SGE_) consequence (OR_ALL_ 10.79 (3.31-35.16)). Carriers of variants with an intermediate function (INT_SGE_) consequence also had, on average, a higher risk than carriers of FUNC_SGE_ variants (OR_ALL_ 3.17 (0.32-31.15)) though the number of INT_SGE_ carriers was small (total n=6). Since the BRCA1 SGE experiment specifically targeted the domain-coding regions of the gene, only four variants outside of the domains were scored. Thus, all *BRCA1* missense variants were assigned to one of four potential risk levels, with SGE score prioritized where available: INT_SGE_/LOF_SGE_; RING/BRCT + Helix-high (SGE score missing); RING/BRCT + Helix-low (SGE score missing); or FUNC_SGE_ or carriers of variants outside of the domains. Compared with non-carriers, there was increased risk for carriers of variants in the INT_SGE_/LOF_SGE_ category (OR 7.22 (2.48-21.01), p=0.00029) and in the RING/BRCT + Helix-high category (OR 5.35 (2.48-11.57), p=2.0×10^−5^; see Additional File 2: Additional Table 5). In a mixture model in which the OR for risk associated missense variants was constrained to that for PTVs (OR 10.69 (7.97-14.33)), the estimated proportions of risk associated variants in the INT_SGE_ /LOF_SGE_ and the RING/BRCT + Helix-high risk categories were 75% (24%-97%) and 51% (6%-94%), respectively (Additional File 2: Additional Table 5). The SGE LR model and SGE mixture model were similarly good fits to the data (2 x log-likelihood difference = 0.12) and both were better fits to the data compared to the Helix-only models (LR models 2 x log-likelihood difference = 3.40, mixture models 2 x log-likelihood difference = 3.58).

### BRCA2

The analysis of *BRCA2* missense variants included 5,467 carriers of 1,425 unique variants. Align-GVGD (p_ALL_=0.0072), BayesDel (p_ALL_=0.059), CADD (p_ALL_=0.036) and Helix (p_ALL_=0.0016) scores were associated with risk for carriers of BRCA2 missense variants (see Additional File 1: Additional Table 3). Risks did not differ by protein domain (p_ALL_=0.91). Compared with non-carriers, carriers of Helix-high variants had a modestly increased risk of breast cancer (OR 1.28 (0.96-1.70), p=0.087; p_ALL_=0.020) whereas carriers of a Helix-low variant had no increased risk (OR 0.98 (0.92-1.04), p=0.47; p_ALL_=0.40; Table 1, Figure 1c, Figure 2c). Under a mixture model in which risk associated missense variants conferred the same risk as PTVs (OR 5.87 (4.75-7.24)), an estimated 11% (4%-25%) of the Helix-high variants were associated with risk, compared with <0.1% of Helix-low variants (Table 1, Figure 1c, Figure 2c). A model that allowed the OR for missense variants to differ from that of PTVs did not converge. The constrained mixture model was a better fit to the data than the logistic regression model (2 x log-likelihood difference = 6.38).

Twelve *BRCA2* variants would be classified as (likely) pathogenic according to ENIGMA guidelines or ClinVar (see Additional File 2: Additional Table 4). In aggregate, the relative risk estimate for these variants was similar to that for PTVs (OR 8.91 (2.61-30.42), p=4.8×10^−4^). Ten of these variants were categorized as Helix-high and two as Helix-low. Two of the variants categorized as (likely) pathogenic and Helix-high were observed in controls only (see Additional File 2: Additional Table 4). After excluding the (likely) pathogenic variants from the LR model, there remained no increased risk associated with variants classified as Helix-high (OR 0.60 (0.27-1.34)).

### CHEK2

The analysis of *CHEK2* missense variants included 1,552 carriers of 325 unique variants. In the carrier-only analysis, BayesDel (p_ALL_=0.0091), CADD (p_ALL_=0.0073), Helix (p_ALL_=0.0021) and REVEL (p_ALL_=0.016) scores were associated with risk (see Additional File 1: Additional Table 3). Compared with non-carriers, carriers of a Helix-high variant had a larger increased risk (OR 1.73 (1.42-2.11), p=4.7×10^−8^) than carriers of Helix-low variants, but the latter were also associated with an increased risk (OR 1.26 (1.08-1.46), p=0.0025; see Table 1, Figure 1d, Figure 2d). There was no significant association with protein domain (p_ALL_=0.98).

In the mixture model analysis, the constrained model in which risk associated missense variants conferred the same risk as PTVs could be rejected (p=0.027). Under the best fitting model, the OR for missense variants was 1.75 (1.47-2.08), with 95% (86%-98%) of Helix-high variants and 33% (25%-43%) of Helix-low variants being risk associated (see Table 1, Figure 1d, Figure 2d). The mixture model was a similar fit to the LR model (2 x log-likelihood difference = 0.52). We also explored mixture models with two levels of risk variant: one with an OR equal to that of PTVs and another conferring a lower risk compared to that of PTVs. The two-level model fitted slightly better in the full training dataset (2 x log-likelihood difference = 1.10) but not in the population-based studies (two-level model converged to the one-level model).

Two variants, c.470T>G (p.Ile157Ser) and c.433C>T (p.Arg145Trp), were listed as (likely) pathogenic on ClinVar; both variants have high Helix scores but the number of carriers in our population-based sample was too small to evaluate their association with risk (see Additional File 2: Additional Table 4). One rare variant, c.349A>G (p.Arg117Gly), was previously identified as risk associated in BCAC samples, as part of the OncoArray genome-wide association study (GWAS) project (26). In the current dataset, this variant, which is in the Helix-high category, had an OR 2.69 (1.46-4.94). After excluding the BCAC GWAS samples from the current dataset, the OR was 3.40 (1.52-7.61). Excluding c.349A>G from the LR model did not change the overall relative risk associated with the Helix-high category (OR 1.64 (1.33-2.02)).

### PALB2

The analysis of *PALB2* missense variants included 1,659 carriers of 472 unique variants. We found no overall evidence of risk associated with missense variants in *PALB2* (OR 0.95 (0.85-1.06), p=0.34; p_ALL_=0.98). In the carrier-only analysis, CADD was the only score associated with risk (p_ALL_=0.020; see Additional File 1: Additional Table 3); however, there was no significant difference in risk between CADD quintiles (p_ALL_=0.16). There was no evidence for a difference in risk for carriers of variants inside any protein domain versus those outside (p_ALL_=0.25). In a mixture model in which the missense variant risk was constrained to that for PTVs (OR 4.87 (3.50-6.77)), the estimated proportion of risk associated variants was 0.011% (95% CI 0%-88%; Table 1, Figure 1e, Figure 2e). The log-likelihoods for the mixture model and logistic regression model were similar (2 x log-likelihood difference = 1.01).

Three (likely) pathogenic variants were listed on ClinVar but none of these were present in our samples. Another variant, c.104T>C (p.Leu35Pro), has been suggested to be pathogenic based on evidence from one family and tumor genomic analysis (28), but this variant was also not found in our samples.

### FREQUENCY ANALYSIS

In the analysis of variant frequency (up to 0.5%) there was no association between risk and frequency, either on a continuous scale or as the difference in risk between rare (<0.1%) and more common (0.1%-0.5%) variant carriers, for *ATM, BRCA2, CHEK2 or PALB2* (see Additional File 2: Additional Table 6). For *BRCA1*, we found frequency inversely associated with risk (continuous p_ALL_=0.022) and a significantly higher risk for rare (<0.1%) compared with more common (0.1%-0.5%) variants (p_ALL_=0.0066). However, after adjusting for the Helix and domain risk groups, neither of these associations remained statistically significant (p_ALL_=0.36 and p_ALL_=0.39, respectively). We evaluated the risks for individual missense variants with frequency between 0.1% and 0.5% (see Additional File 2: Additional Table 7). In *ATM*, one variant, c.5312G>A (p.Arg1771Lys), was associated with a modest increase in risk (OR=1.33 (0.92-1.93), p=0.13; p_ALL_=0.0070). In *BRCA1*, one variant, c.2521C>T (p.Arg841Trp), was associated with a decreased risk of breast cancer (OR 0.67 (0.52-0.87), p=0.0027). Two previously-reported variants in *CHEK2* were identified: c.470T>C (p.Ile157Thr) and c.538C>T (p.Arg180Cys) (29). c.470T>C was associated with an OR of 1.24 (1.09-1.42), consistent with the estimate for the Helix-low risk category, while c.538C>T was associated with a higher OR 1.44 (1.12-1.84). No *BRCA2* or *PALB2* missense variants were individually associated with risk.

### MODEL VALIDATION

We evaluated the calibration of the best fitting models from the training set, for each gene, in the validation set: these included the LR models, the mixture model using the estimated proportions (α) from the training set, and the mixture model using the posterior probabilities derived from the training set. For each gene and each model, carriers of variants in the predicted risk groups were associated with an increased risk, and there were no differences between the observed and predicted ORs (see Additional File 2: Additional Table 8 and Additional Figures 2-6). *In-silico* scores, likelihood ratios and posterior probabilities for every variant included in the population training dataset are given in Additional File 1: Additional Tables 9-13.

Using a composite five gene model, we estimated ORs for eleven risk categories (Figure 3). 184 samples carried a missense variant in more than one of the five genes and were excluded from this analysis. Four categories were significantly associated with an increased risk relative to non-carriers, consistent with the estimates derived from the training set: *ATM* FAT/PIK + CADD5 (OR 1.76 (1.16-2.68), p=0.0078), *CHEK2* Helix-low (OR 1.40 (1.04-1.88), p=0.025), *CHEK2* Helix-high (OR=1.89 (1.27-2.81), p=0.0017) and *BRCA1* within domain and Helix-high (OR 4.44 (1.45-13.59), p=0.0089) risk groups. The OR estimate for *BRCA2* Helix-high variant carriers was higher than that in the training dataset, but the confidence interval was considerably wider (OR 1.54 (0.88-2.68)). As predicted, variants in the remaining categories were not associated with risk.

**Figure 3.**
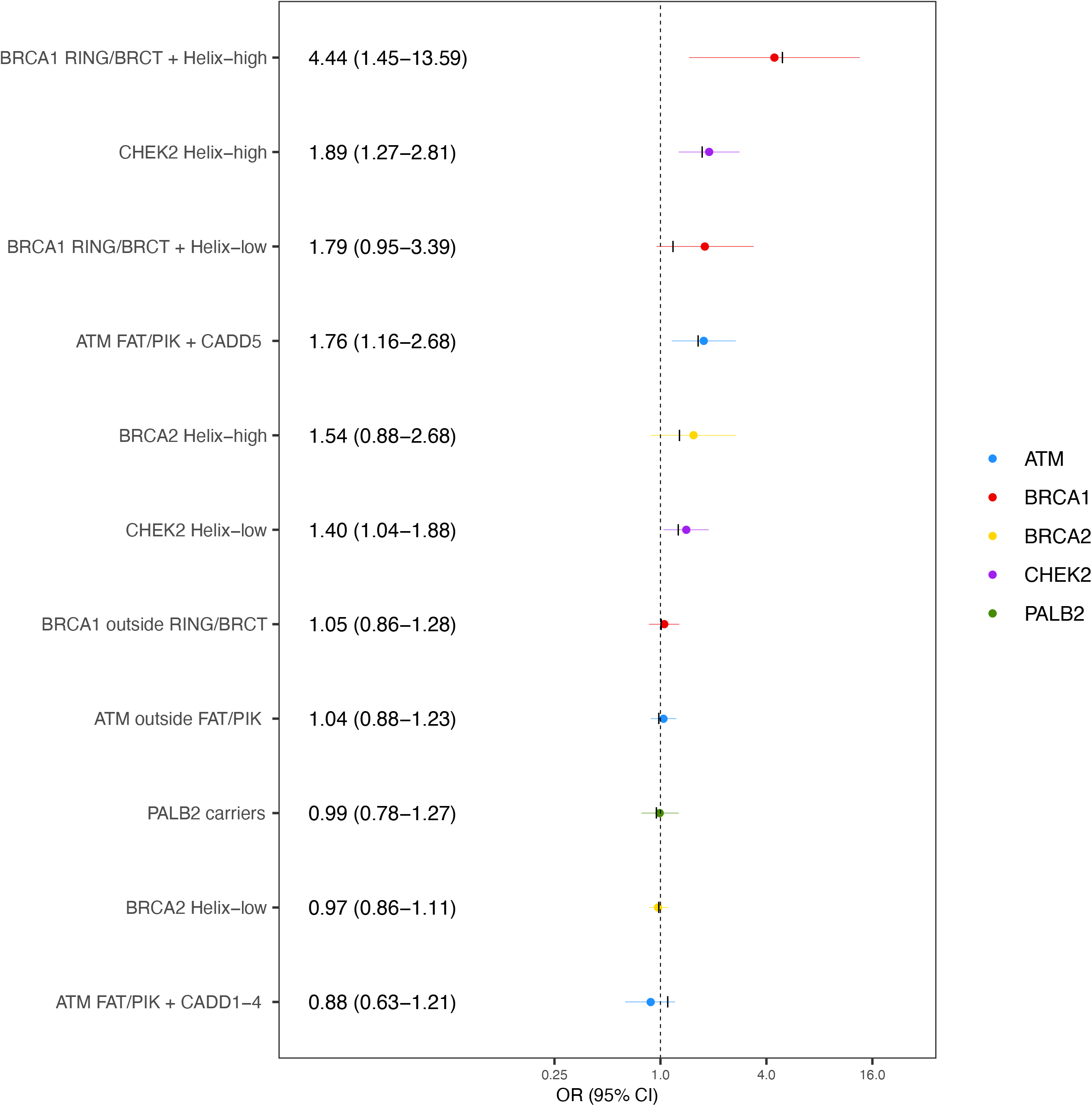
Breast cancer risk estimates from composite gene model in validation samples. **Figure 3 Legend** Black marks indicate corresponding ORs from training models. Risk categories: ATM FAT/PIK + CADD5: *ATM* variants lying within the FAT or PI3K/PI4K protein domains with CADD score in fifth quintile; ATM FAT/PIK+CADD1-4: *ATM* variants lying within the FAT or PI3K/PI4K protein domains with CADD score in any of first four quintiles; ATM outside FAT/PIK: variants lying outside the FAT and PI3K/PI4K protein domains; BRCA1 RING/BRCT + Helix-high: *BRCA1* variants lying within the RING or BRCT domains with a high Helix score; BRCA1 RING/BRCT + Helix-low: *BRCA1* variants lying with the RING or BRCT domains with a low Helix score; BRCA1 outside RING/BRCT: *BRCA1* variants lying outside the RING and BRCT domains; BRCA2 Helix-high: *BRCA2* variants with a high Helix score; BRCA2 Helix-low: *BRCA2* variants with a low Helix score; CHEK2 Helix-high: *CHEK2* variants with a high Helix score; CHEK2 Helix-low: *CHEK2* variants with a low Helix score; PALB2 carriers: carriers of any missense variant in *PALB2*

## Discussion

To date, the risks associated with missense variants in breast cancer predisposition genes have been largely unclear. In this study of over 112,000 women, we were able to use a range of *in silico* scores produced by statistical algorithms and knowledge of functional protein domains to determine the risks associated with subsets of rare missense variants. We identified groups of missense variants conferring increased risks of breast cancer in *ATM, BRCA1, BRCA2* and *CHEK2*, but not in *PALB2*. Under the best fitting mixture models, for *ATM, BRCA1* and *BRCA2*, a small proportion of rare missense variants were associated with risks comparable to those for PTVs. In contrast, for *CHEK2*, a high proportion of *CHEK2* missense variants were risk associated and the estimated risk was markedly lower than that associated with PTVs. In *PALB2*, the evidence for association was weak; the mixture model analysis indicated that the proportion of missense variants associated with a high risk is likely to be very small.

We used five *in silico* scores to predict the pathogenicity of individual variants. Helix, BayesDel and CADD were all predictive for the four genes for which we were able to identify subsets of risk-associated variants; Helix was most predictive for *BRCA1, BRCA2* and *CHEK2* while CADD outperformed all the other scores for *ATM*. In addition to the *in silico* scores, we also tested the BRCA1 SGE functional assay score. We found that the SGE score slightly improved the performance of the model for predicting risk for *BRCA1* missense variant carriers, compared with the Helix-only model. Variants categorized by SGE as disruptive to function, or lying within a protein domain and scored high by Helix, were strongly associated with increased risk. Under the mixture model, the proportions of risk-associated variants were also high, although the confidence intervals for the proportion of associated variants were wide. It is notable that 11 of the 31 variants in these categories have previously been identified as (likely) pathogenic by ClinVar and/or ENIGMA. Similarly, in *ATM*, the risk conferred by missense variants was confined to specific protein-coding domains, namely the FAT and PIK domains, consistent with previous studies (5). Variants within these domains could be further distinguished using the CADD score; variants in the top quintile were associated with risk whereas variants in the first four quintiles were not. In a mixture model, 54% of variants in the top CADD quintile were estimated to be associated with risk. One variant in this group, c.7271T>G (p.Val2424Gly), has been previously reported as a breast cancer risk variant but the OR estimate for this variant, 1.63 (0.56-4.73), was markedly lower than previously estimated (relative risks ranging from 8.0-12.7)(24-26). The reasons for this difference are unclear but might be due, in part, to previous studies oversampling for cases with a family history of breast cancer. In *BRCA2*, a small number of variants were categorized as deleterious by Helix and showed an overall association with risk in our study (OR 1.28 (0.96-1.70), p=0.087; p_ALL_=0.020). Under the best fitting mixture model, a small proportion (11%) of these variants were estimated to be associated with risk equivalent to that of PTVs (OR 5.87 (4.75-7.24)).

The results for *CHEK2* were in marked contrast to those for *BRCA1, BRCA2* and *ATM*. In the best fitting mixture model, the proportion of associated variants was high, and the estimated risk was clearly lower than for PTVs. A model in which there were two levels of risk, with the higher level equal to the PTV risk, fitted slightly better in the full training dataset but not in the population-only training studies. In addition, however, three individual *CHEK2* variants were associated with differing levels of risk: c.470T>C (p.Ile157Thr) OR 1.24 (1.09-1.42); c.538C>T (p.Arg180Cys) OR 1.44 (1.12-1.84); and c.349A>G (p.Arg117Gly) OR 2.69 (1.46-4.94). The c.470T>C variant was too common to be included in the main analyses, possibly explaining why the heterogeneity in risk was not readily detectable by the mixture models; however, the confidence interval for c.470T>C from the individual level analysis did not include the LR and mixture model OR estimates of 1.73 and 1.75, respectively, for the risk-associated variants. Taken together, these observations suggest that there is substantial variation in risk associated with *CHEK2* missense variants.

Under the best fitting mixture model, approximately 7% of all rare missense variants in *ATM* were associated with similar risk to that of PTVs. The estimated carrier frequency of pathogenic missense variants in *ATM* was 0.0030, or approximately 89% of the PTV frequency. The corresponding proportion of associated rare missense variants for *BRCA1* and *BRCA2* was 2% and 0.6%, with an estimated carrier frequency of 0.00026 (∼18%) and 0.00028 (∼10%), respectively. Thus, missense variants add modestly to the contribution of *BRCA1* and *BRCA2* variants to breast cancer incidence, but make a relatively more substantial contribution for *ATM*. For *CHEK2*, approximately 60% of rare missense variants were risk associated and the estimated carrier frequency of pathogenic missense variants in *CHEK2* was comparable to the frequency of PTVs. The predicted proportion of breast cancer cases possessing pathogenic germline missense variants in these genes is approximately 0.6%, 0.3%, 0.2% and 1.3% for *ATM, BRCA1, BRCA2* and *CHEK2*, respectively.

The task of identifying which specific individual missense variants are risk associated is a complex one and is difficult to resolve fully even with a large dataset, since most variants are rare and there are many possible models to consider. Despite the size of our study, it was difficult to distinguish, for any gene, between the LR models (in which all variants in a given category confer a given risk) and the mixture models (in which all risk-associated variants confer the same risk, but the proportion that are associated varies by category). This difficulty arises because the number of carriers for individual variants is small, and as a result the estimated risk of pathogenic missense variants and probability of pathogenicity (α) are strongly confounded. Further, selecting the best models and estimating the risks based on these models is likely to result in overfitting and biased risk estimates. In order to strengthen the validity of our findings, we used a training-validation study design. We were able to replicate the predicted OR estimates in the validation dataset, suggesting that any bias due to overfitting was small. Nevertheless, the validation dataset was relatively small, so further validation of the best models reported here in large independent datasets is critical.

Ultimately, high-throughput functional assays may provide more precise definitions of risk categories. The analysis of the SGE scores for *BRCA1* suggests that this approach should be useful, although the scores were highly concordant with the best *in-silico* score in this case. This study provides an evidence-based starting point to select candidate variants for future functional assays. Ultimately, however, larger population-based epidemiological datasets will be required to provide more precise risk estimates.

## Conclusions

This study confirms that subsets of missense variants in established breast cancer susceptibility genes are associated with increased risks of the disease and provides estimates of relative risks for those subsets, as well as probabilities for association with risk at the variant level. The pattern of risk varies substantially by gene. Accurately and precisely defining these risks is critical to the counselling and management of women in whom these variants are identified.

## Supporting information

Additional Figures 1-6, Additional Tables 2, 4-8, and Additional Note (funding and acknowledgements)

## Data Availability

Summary level genotype data are available via http://bcac.ccge.medschl.cam.ac.uk and in Supplementary Tables 9-13. Individual level data are available via the BCAC Data Access Co-ordinating Committee

## List of abbreviations

PTVs: Protein truncating variants
VUS: Variants of uncertain significance
BCAC: Breast Cancer Association Consortium
VEP: Ensembl Variant Effect Predictor
CADD: Combined Annotation Dependent Depletion
REVEL: Rare Exome Variant Ensembl Learner
SGE: Saturation Genome Editing
LR: Logistic regression
OR: Odds ratio
EM: Expectation-maximization
PP: Posterior probability
Q5: Fifth quntile
LOF_SGE_: Loss of function
FUNC_SGE_: Functional
INT_SGE_: Intermediate function
GWAS: Genome wide association study

## Declerations

### ETHICS APPROVAL AND CONSENT TO PARTICPATE

Approval to use the BCAC data included in this study was given by the Data Access Co-ordinating Committee (concept #672). All contributing BCAC studies were approved by the relevant ethical review boards, details of which can be found in Additional Table 14.

### CONSENT FOR PUBLICATION

Not applicable

### AVAILABILITY OF DATA AND MATERIALS

Summary level genotype data are available via http://bcac.ccge.medschl.cam.ac.uk and in Additional Tables 9-13. Individual level data are available via the BCAC Data Access Co-ordinating Committee

### FUNDING

The sequencing and analysis for this project was funded by the European Union’s Horizon 2020 Research and Innovation Programme (BRIDGES: grant number 634935) and the Wellcome Trust [grant no: v203477/Z/16/Z]. BCAC co-ordination was additionally funded by the European Union’s Horizon 2020 Research and Innovation Programme (BRIDGES: grant number 634935, BCAST: grant number 633784) and by Cancer Research UK [C1287/A16563]. Study specific funding is given in the Additional Note.

## AUTHORS’ CONTRIBUTIONS

DFE, PD and SWT conceived the study and obtained funding. LD, SC, JMA performed the statistical and bioinformatics analysis. MTP and CF performed variant annotation analysis. SMH and BV developed the Helix algorithm. A G-N, JB, AMD, CL and AK led the laboratory analysis. LD and DFE drafted the manuscript. ABS, MPGV, PD, SWT, MKS, MdlH and AMD revised the manuscript. All other authors generated study-specific data. All authors read and approved the final manuscript.

## ACKNOWLEDGEMENTS

*kConFab Investigators*

Adrienne Sexton, Alex Dobrovic, Alice Christian, Alison Trainer, Allan Spigelman, Andrew Fellows, Andrew Shelling, Anna De Fazio, Anneke Blackburn, Ashley Crook, Bettina Meiser, Briony Patterson, Christine Clarke, Christobel Saunders, Clare Hunt, Clare Scott, David Amor, David Gallego Ortega, Deb Marsh, Edward Edkins, Elizabeth Salisbury, Eric Haan, Finlay Macrea, Gelareh Farshid, Geoff Lindeman, Georgia Trench, Graham Mann, Graham Giles, Grantley Gill, Heather Thorne, Ian Campbell, Ian Hickie, Liz Caldon, Ingrid Winship, James Cui, James Flanagan, James Kollias, Jane Visvader, Jennifer Stone, Jessica Taylor, Jo Burke, Jodi Saunus, John Forbes, John Hopper, Jonathan Beesley, Judy Kirk, Juliet French, Kathy Tucker, Kathy Wu, Kelly Phillips, Laura Forrest, Lara Lipton, Leslie Andrews, Lizz Lobb, Logan Walker, Maira Kentwell, Mandy Spurdle, Margaret Cummings, Margaret Gleeson, Marion Harris, Mark Jenkins, Mary Anne Young, Martin Delatycki, Mathew Wallis, Matthew Burgess, Melissa Brown, Melissa Southey, Michael Bogwitz, Michael Field, Michael Friedlander, Michael Gattas, Mona Saleh, Morteza Aghmesheh, Nick Hayward, Nick Pachter, Paul Cohen, Pascal Duijf, Paul James, Pete Simpson, Peter Fong, Phyllis Butow, Rachael Williams, Rick Kefford, Rodney Scott, Roger Milne, Rosemary Balleine, Sarah – Jane Dawson, Sheau Lok, Shona O’Connell, Sian Greening, Sophie Nightingale, Stacey Edwards, Stephen Fox, Sue-Anne McLachlan, Sunil Lakhani, Tracy Dudding, Yoland Antill.

*NBCS Collaborators*

Kristine K. Sahlberg, Anne-Lise Børresen-Dale, Inger Torhild Gram, Olav Engebråten, Bjørn Naume, Jürgen Geisler, OSBREAC, Grethe I. Grenaker Alnæs.

*SGBCC Investigators*

Swee Ho Lim, Ern Yu Tan, Benita Kiat Tee Tan, Su-Ming Tan, Veronique Kiak Mien Tan, Ching Wan Chan, Siau-Wei Tang, Celene Wei Qi Ng, Geok Hoon Lim, Jinnie Siyan Pang, Jung Ah Lee, Patrick Mun Yew Chan, Juliana Chen, Sarah Qinghui Lu, Yirong Sim, Wei Sean Yong, Preetha Madhukumar, Fuh Yong Wong, Joanne Yuen Yie Ngeow, Tira Jing Ying Tan, Wai Peng Lee, Chi Wei Mok, Chin Mui Seah, Linda Tan, E Shyong Tai, Xueling Sim, Peh Joo Ho, Alexis Jiaying Khng.

## Additional Files

Additional File 1: Excel spreadsheet containing Additional Table 1, Additional Table 3 and Additional Tables 9-14.

Additional File 2: PDF document containing Additional Figures 1-6, Additional Table 2, Additional Tables 4-8 and an Additional Note containing funding and acknowledgement details.

