## Additional Figures 1-6, Additional Tables 2, 4-8, and Additional Note (funding and acknowledgements) for "Breast cancer risks associated with missense variants in breast cancer susceptibility genes"

Additional Figure 1: Flow diagram of statistical analysis. Training analyses in blue shaded area and validation analyses in yellow shaded area. Boxes outlined in orange show analyses based on population samples only.

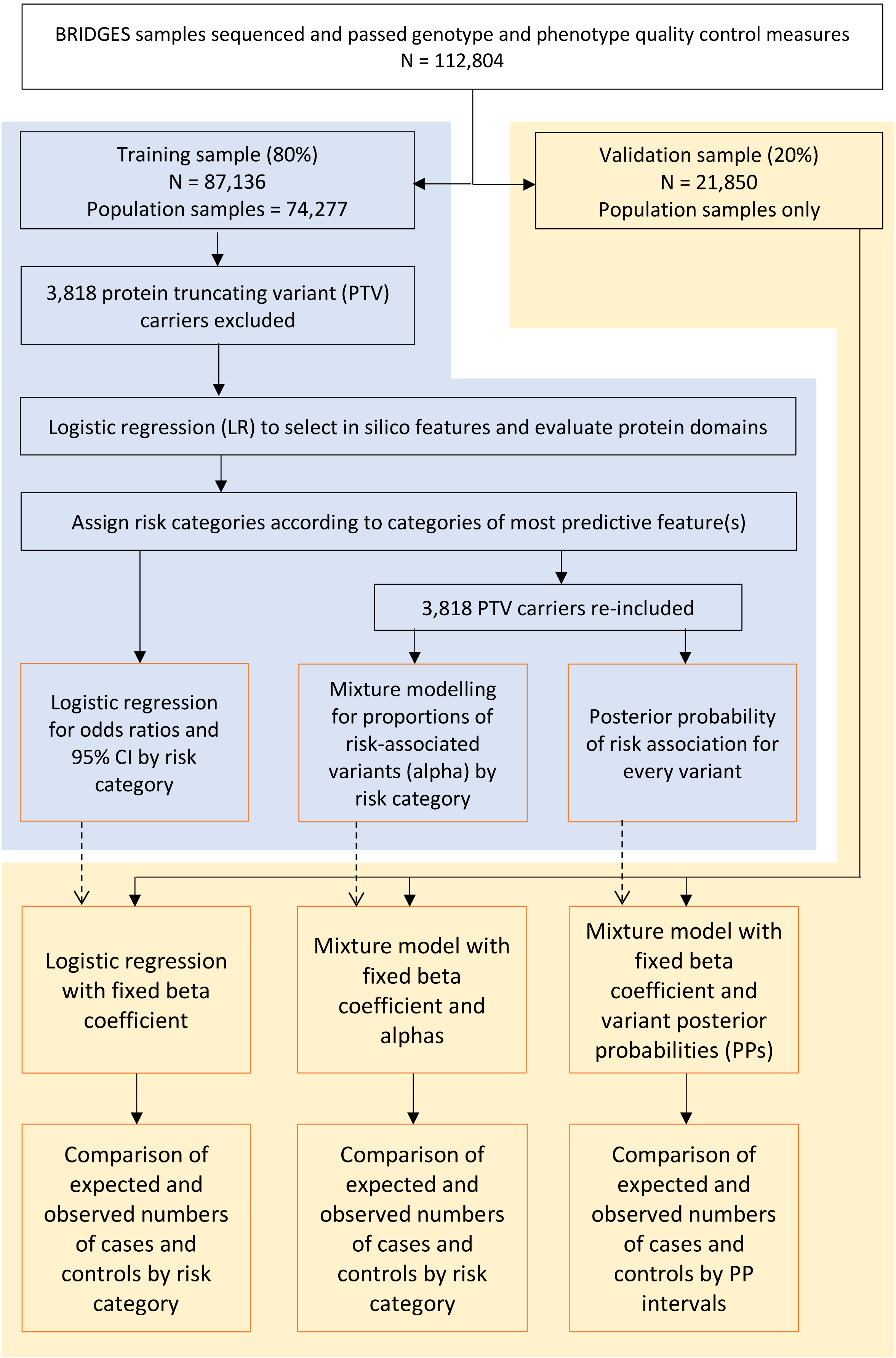

Additional Figure 2: Plot of expected versus observed ORs for carriers of *ATM* missense variants in the test dataset, by risk category under the logistic regression model with fixed beta; by risk category under the mixture model with fixed alphas and beta; by intervals of variant posterior probabilities (PPs) under the mixture model with fixed PPs.

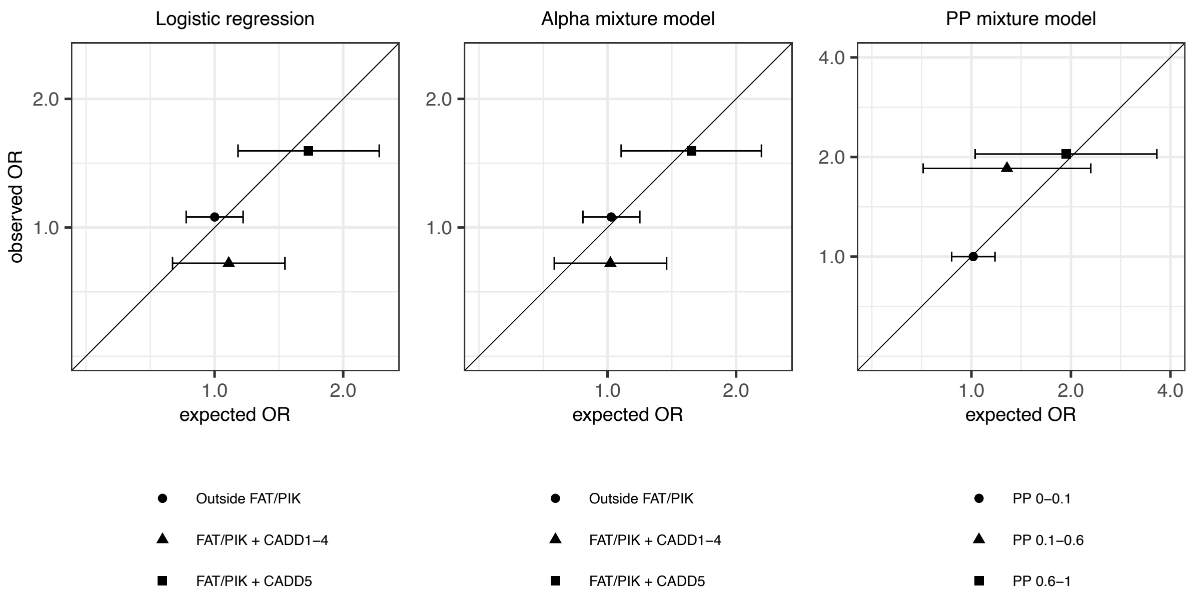

Additional Figure 3: Plot of expected versus observed ORs for carriers of *BRCA1* missense variants in the test dataset, by risk category under the logistic regression model with fixed beta; by risk category under the mixture model with fixed alphas and beta; by intervals of variant posterior probabilities (PPs) under the mixture model with fixed PPs.

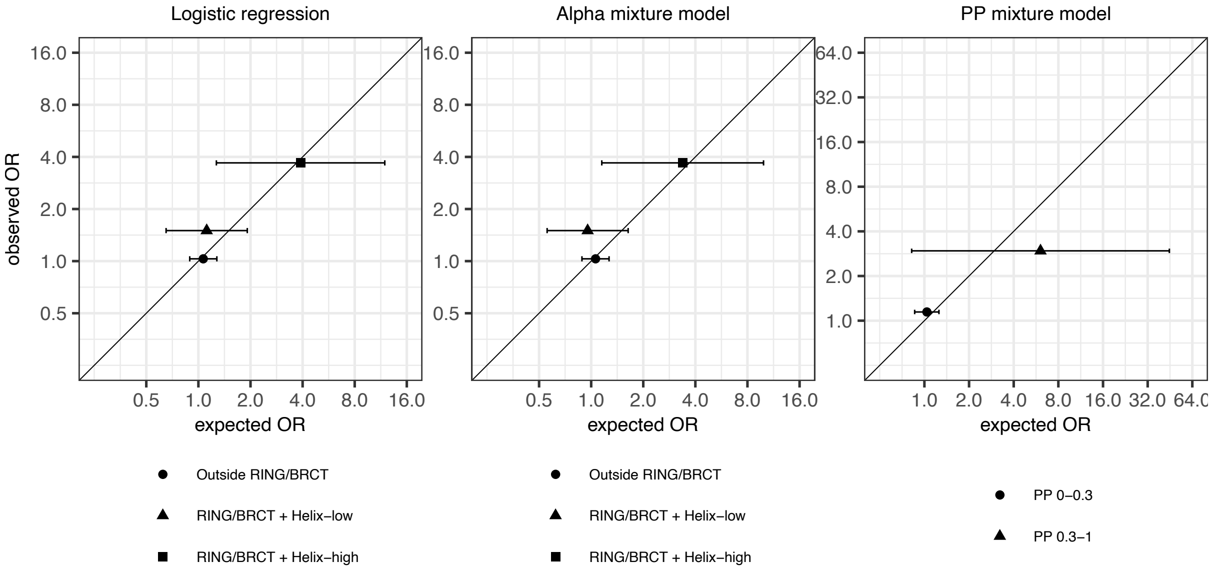

Additional Figure 4: Plot of expected versus observed ORs for carriers of *BRCA2* missense variants in the test dataset, by risk category under the logistic regression model with fixed beta; by risk category under the mixture model with fixed alphas and beta; by intervals of variant posterior probabilities (PPs) under the mixture model with fixed PPs.

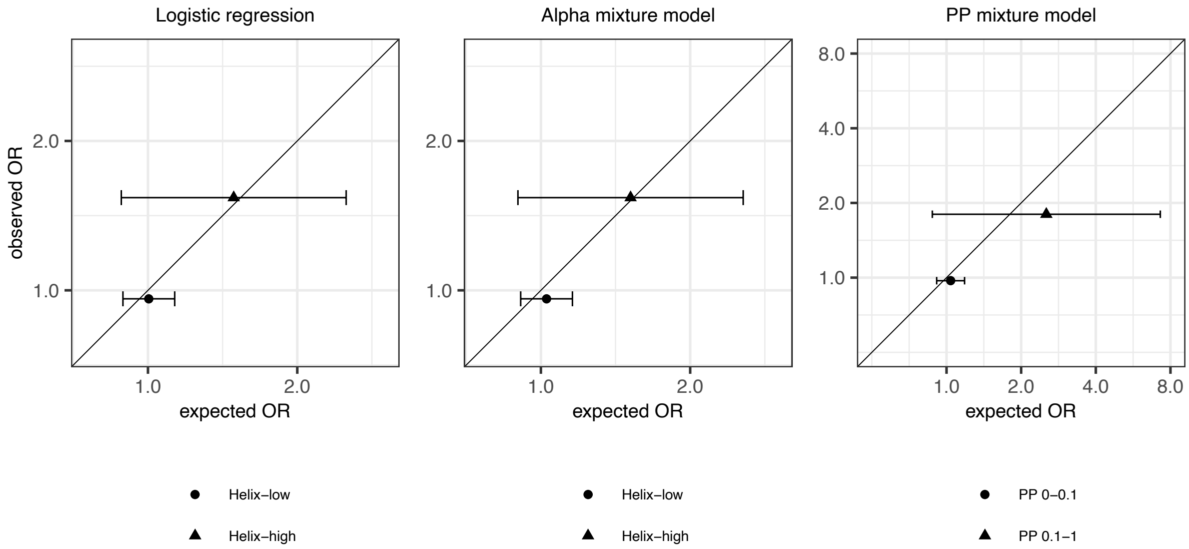

Additional Figure 5: Plot of expected versus observed ORs for carriers of *CHEK2* missense variants in the test dataset, by risk category under the logistic regression model with fixed beta; by risk category under the mixture model with fixed alphas and beta; by intervals of variant posterior probabilities (PPs) under the mixture model with fixed PPs.

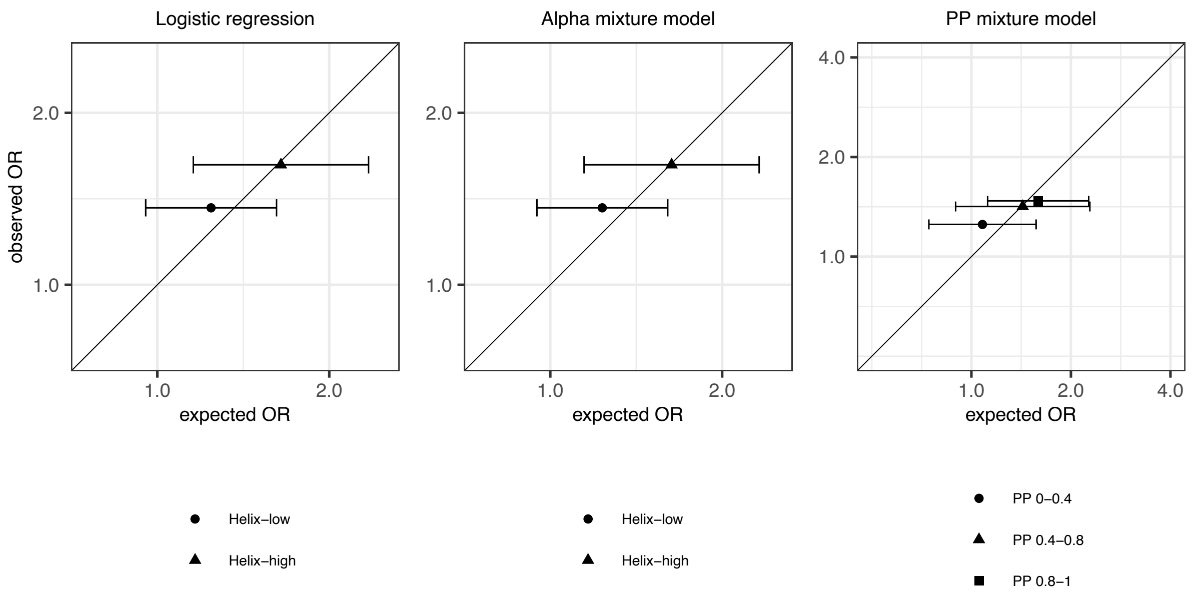

Additional Figure 6: Plot of expected versus observed ORs for carriers of *PALB2* missense variants in the test dataset, by risk category under the logistic regression model with fixed beta; by risk category under the mixture model with fixed alphas and beta; by intervals of variant posterior probabilities (PPs) under the mixture model with fixed PPs.

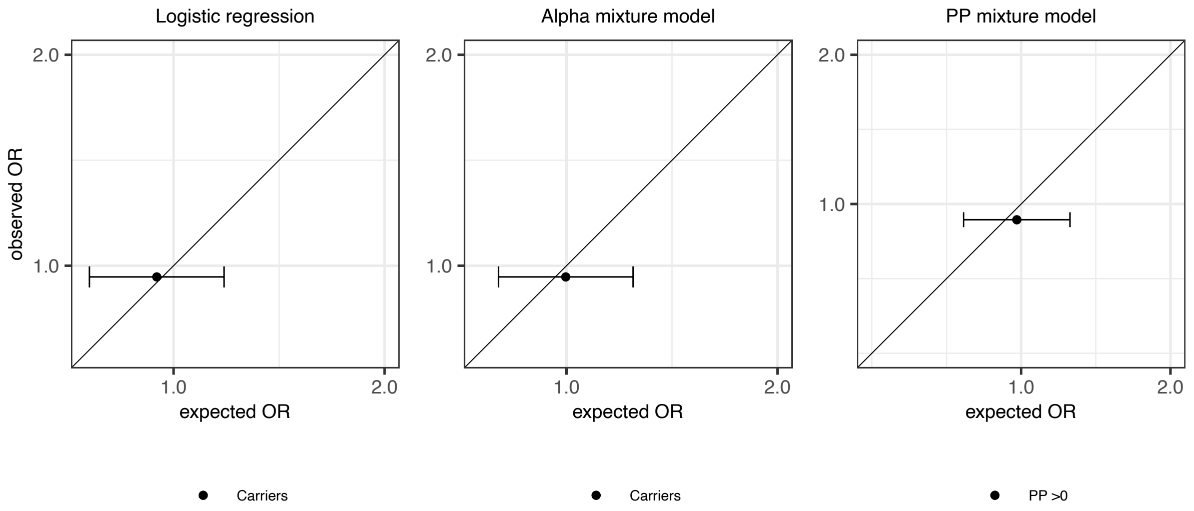

Additional Table 2: Functional protein domain definitions

| Gene | Domain | Amino acids | Reference |
| --- | --- | --- | --- |
| *ATM* | FRAP, ATM, TRRAP (FAT) | 1966-2566 | Lau, W.C. et al, 2016. Structure of the human dimeric ATM kinase. *Cell Cycle*, *15*(8), pp.1117-1124. |
| | PI3/PI4-Kinase (PIK) | 2614-2960 | |
| | FRAP, ATM, TRRAP C-terminal (FAT-C) | 3025-3056 | |
| *BRCA1* | RING finger | 1-101 | ENIGMA: Evidence-based Network for the Interpretation of Germline Mutant Alleles, available at https://enigmaconsortium.org[a] |
| | BRCA1 C-terminal (BRCT) I-II | 1642-1863 | |
| *BRCA2* | PALB2 binding | 10-40 | ENIGMA: Evidence-based Network for the Interpretation of Germline Mutant Alleles, available at https://enigmaconsortium.org[a] |
| | DNA binding | 2481-3186 | |
| *CHEK2* | SQ/TQ | 19-69 | Cai, Z. et al, 2009. Structure and activation mechanism of the CHK2 DNA damage checkpoint kinase. *Molecular cell*, *35*(6), pp.818-829. |
| | Forkhead-associated (FHA) | 92-205 | |
| | Kinase | 212-501 | |
| *PALB2* | Coiled-coil | 9-44 | Rodrigue, A. et al, 2019. A global functional analysis of missense mutations reveals two major hotspots in the PALB2 tumor suppressor. *Nucleic acids research*, *47*(20), pp.10662-10677.<br>Boonen, R.A. et al, 2019. Functional analysis of genetic variants in the high-risk breast cancer susceptibility gene PALB2. *Nature communications*, *10*(1), pp.1-15. |
| | Chromatin-association motif (ChAM) | 394-446 | |
| | DNA binding | 611-764 | |
| | WD40 | 853-1186 | |

[a]drawn from ENIGMA ClinGen External Expert Panel Rules Version 2.5.1, describing conserved domains/motifs known to harbour clinically important missense alterations

Additional Table 4: BRIDGES missense variants classified as (Likely) Pathogenic on ClinVar (*ATM*, *CHEK2*, *PALB2, BRCA1, BRCA2*) or by ENIGMA expert guidelines (*BRCA1*, *BRCA2*).

| Gene | Variant | N cases[a] | N controls[a] |
|---|---|---|---|
| *ATM* | c.875C>T (p.Pro292Leu) | 1 | 0 |
| *ATM* | c.2849T>G (p.Leu950Arg) | 1 | 0 |
| *ATM* | c.3848T>C (p.Leu1283Pro) | 0 | 1 |
| *ATM* | c.6200C>A (p.Ala2067Asp) | 1 | 0 |
| *ATM* | c.6679C>T (p.Arg2227Cys) | 5 | 0 |
| *ATM* | c.7271T>G (p.Val2424Gly) | 12 | 6 |
| *ATM* | c.7570G>C (p.Ala2524Pro) | 4 | 0 |
| *ATM* | c.8122G>A (p.Asp2708Asn) | 2 | 0 |
| *ATM* | c.8147T>C (p.Val2716Ala) | 9 | 5 |
| *ATM* | c.8494C>T (p.Arg2832Cys) | 8 | 2 |
| *ATM* | c.8546G>C (p.Arg2849Pro) | 0 | 1 |
| *ATM* | c.9022C>T (p.Arg3008Cys) | 4 | 1 |
| *ATM* | c.9023G>A (p.Arg3008His) | 3 | 1 |
| | *Estimate for aggregated variants* | *OR (95% CI)* | *P-value* |
| | *All samples* | *2.74 (1.55-4.86)* | *0.00053* |
| | *Population samples* | *1.85 (0.98-3.50)* | *0.060* |
| *BRCA1* | c.5339T>C (p.Leu1780Pro) | 9 | 2 |
| *BRCA1* | c.5216A>G (p.Asp1739Gly) | 1 | 0 |
| *BRCA1* | c.5213G>A (p.Gly1738Glu) | 3 | 0 |
| *BRCA1* | c.5207T>C (p.Val1736Ala) | 1 | 0 |
| *BRCA1* | c.5095C>T (p.Arg1699Trp) | 2 | 0 |
| *BRCA1* | c.5089T>C (p.Cys1697Arg) | 2 | 0 |
| *BRCA1* | c.5072C>T (p.Thr1691Ile) | 2 | 0 |
| *BRCA1* | c.5057A>G (p.His1686Arg) | 4 | 0 |
| *BRCA1* | c.4964C>T (p.Ser1655Phe) | 1 | 0 |
| *BRCA1* | c.181T>G (p.Cys61Gly) | 28 | 3 |
| *BRCA1* | c.181T>C (p.Cys61Arg) | 1 | 0 |
| *BRCA1* | c.130T>A (p.Cys44Ser) | 2 | 1 |
| *BRCA1* | c.53T>C (p.Met18Thr) | 4 | 0 |
| | *Estimate for aggregated variants* | *OR (95% CI)* | *P-value* |
| | *All samples* | 9.97 (4.29-23.17) | $9.2 \times 10^{-8}$ |
| | *Population samples* | 16.68 (5.16-53.94) | $2.6 \times 10^{-6}$ |
| *BRCA2* | c.7529T>C (p.Leu2510Pro) | 2 | 1 |
| *BRCA2* | c.7879A>T (p.Ile2627Phe) | 0 | 1 |
| *BRCA2* | c.7940T>C (p.Leu2647Pro) | 1 | 0 |
| *BRCA2* | c.7958T>C (p.Leu2653Pro) | 0 | 1 |
| *BRCA2* | c.8023A>G (p.Ile2675Val) | 3 | 0 |

| Gene | Variant | N cases[a] | N controls[a] |
|---|---|---|---|
| *BRCA2* | c.8057T>C (p.Leu2686Pro) | 1 | 0 |
| *BRCA2* | c.8167G>C (p.Asp2723His) | 10 | 0 |
| *BRCA2* | c.8243G>A (p.Gly2748Asp) | 3 | 0 |
| *BRCA2* | c.9004G>A (p.Glu3002Lys) | 4 | 0 |
| *BRCA2* | c.9154C>T (p.Arg3052Trp) | 5 | 0 |
| *BRCA2* | c.9226G>A (p.Gly3076Arg) | 3 | 0 |
| *BRCA2* | c.9371A>T (p.Asn3124Ile) | 11 | 1 |
| | *Estimate for aggregated variants* | *OR (95% CI)* | *P-value* |
| | *All samples* | 10.31 (3.65-29.11) | $1.1x10^{-5}$ |
| | *Population samples* | 8.91 (2.61-30.42) | $4.8x10^{-4}$ |
| *CHEK2* | c.470T>G (p.Ile157Ser) | 1 | 0 |
| *CHEK2* | c.433C>T (p.Arg145Trp) | 17 | 7 |

[a] Number of carriers in complete training dataset

Additional Table 5: Breast cancer risk association results from *BRCA1* risk model including SGE score of population samples in the training dataset

| | N | | | Logistic regression model | | | Mixture model | | |
|---|---|---|---|---|---|---|---|---|---|
| Risk group | Variants[a] | Cases | Controls | OR[b] | 95% CI[c] | P-value | Missense OR (95% CI)[d] | α[e] | 95% CI[f] |
| *BRCA1 – SGE model* | | | | Log-likelihood = -48650.44 | | | Log-likelihood = -48650.50 | | |
| Non-carriers | - | 34191 | 37996 | 1 | - | - | | 0 | - |
| Carriers | | | | | | | 10.69 (7.97-14.33)[g] | | |
| $FUNC_{SGE}$ or variant outside RING and BRCT domains | 538 | 878 | 922 | 1.02 | (0.93-1.12) | 0.68 | | $9.4x10^{-4}$ | $(3.2x10^{-5}$-0.027) |
| Variant inside RING or BRCT domain and low Helix score | 20 | 55 | 41 | 1.29 | (0.85-1.96) | 0.22 | | $2.9x10^{-25}$ | NA |
| Variant inside RING or BRCT domain and high Helix score | 3 | 37 | 8 | 5.35 | (2.48-11.57) | $2.0x10^{-5}$ | | 0.51 | (0.062-0.94) |
| $SGE_{INT}$ or $SGE_{LOF}$ | 20 | 24 | 4 | 7.22 | (2.48-21.01) | $2.9x10^{-4}$ | | 0.75 | (0.24-0.97) |

[a] Number of unique missense substitutions in population dataset
[b] Logistic regression odds ratio estimate for missense variant carriers
[c] 95% confidence interval for logistic regression OR estimate for missense variant carriers
[d] Mixture model odds ratio and 95% confidence interval for missense variant carriers
[e] Alpha: estimated proportion of risk associated missense variants
[f] 95% confidence interval for alpha
[g] Missense variant odds ratio constrained to equal odds ratio for protein truncating variants

Additional Table 6: P-values for association of variant frequency up to 0.5% with breast cancer risk in all missense variant carriers in training dataset

| | *P*-value | | |
|---|---|---|---|
| Gene | Continuous frequency | Log-scale | Frequency <0.1% versus frequency 0.1%-0.5% |
| *ATM* | 0.14 | 0.057 | 0.15 |
| *BRCA1* | 0.022 | 0.011 | 0.0066 |
| *BRCA2* | 0.068 | 0.79 | 0.18 |
| *CHEK2* | 0.075 | 0.53 | 0.21 |
| *PALB2* | 0.62 | 0.62 | 0.69 |

Additional Table 7: Breast cancer risk association results for individual missense variants with frequency between 0.1% and 0.5% based on population samples in the training dataset

| Gene | Variant | Case carriers | Control carriers | OR | 95% CI | *P*-value | $P_{ALL}$[a] |
|---|---|---|---|---|---|---|---|
| *ATM* | c.378T>A (p.Asp126Glu) | 46 | 32 | 1.56 | (0.98-2.47) | 0.060 | 0.067 |
| *ATM* | c.998C>T (p.Ser333Phe) | 89 | 94 | 1.11 | (0.82-1.50) | 0.49 | 0.30 |
| *ATM* | c.1229T>C (p.Val410Ala) | 118 | 153 | 0.88 | (0.68-1.13) | 0.31 | 0.32 |
| *ATM* | c.1810C>T (p.Pro604Ser) | 54 | 58 | 1.00 | (0.68-1.48) | 0.99 | 0.34 |
| *ATM* | c.3925G>A (p.Ala1309Thr) | 70 | 110 | 0.73 | (0.53-1.00) | 0.050 | 0.041 |
| *ATM* | c.4138C>T (p.His1380Tyr) | 137 | 159 | 0.98 | (0.77-1.25) | 0.88 | 0.90 |
| *ATM* | c.5071A>C (p.Ser1691Arg) | 162 | 221 | 0.87 | (0.70-1.07) | 0.19 | 0.039 |
| *ATM* | c.5312G>A (p.Arg1771Lys) | 65 | 53 | 1.33 | (0.92-1.93) | 0.13 | 0.0070 |
| *ATM* | c.6067G>A (p.Gly2023Arg) | 120 | 147 | 0.93 | (0.72-1.19) | 0.55 | 0.53 |
| *ATM* | c.6313A>G (p.Arg2105Gly) | 128 | 130 | 1.17 | (0.91-1.50) | 0.22 | 0.10 |
| *BRCA1* | c.4535G>T (p.Ser1512Ile) | 238 | 310 | 0.93 | (0.78-1.11) | 0.43 | 0.45 |
| *BRCA1* | c.2566T>C (p.Tyr856His) | 141 | 138 | 0.98 | (0.77-1.25) | 0.88 | 0.85 |
| *BRCA1* | c.2521C>T (p.Arg841Trp) | 96 | 165 | 0.67 | (0.52-0.87) | 0.0027 | 0.0016 |
| *BRCA2* | c.125A>G (p.Tyr42Cys) | 101 | 109 | 1.03 | (0.78-1.37) | 0.82 | 0.81 |
| *BRCA2* | c.978C>A (p.Ser326Arg) | 95 | 96 | 1.05 | (0.79-1.41) | 0.73 | 0.54 |
| *BRCA2* | c.1151C>T (p.Ser384Phe) | 76 | 97 | 0.87 | (0.63-1.18) | 0.37 | 0.42 |
| *BRCA2* | c.1792A>G (p.Thr598Ala) | 147 | 155 | 1.03 | (0.81-1.30) | 0.80 | 0.54 |
| *BRCA2* | c.5785A>G (p.Ile1929Val) | 79 | 71 | 1.00 | (0.72-1.39) | 0.98 | 0.96 |
| *BRCA2* | c.6100C>T (p.Arg2034Cys) | 301 | 309 | 1.06 | (0.90-1.26) | 0.47 | 0.35 |
| *BRCA2* | c.8149G>T (p.Ala2717Ser) | 98 | 130 | 0.84 | (0.64-1.10) | 0.21 | 0.15 |
| *BRCA2* | c.8182G>A (p.Val2728Ile) | 219 | 208 | 1.25 | (1.02-1.53) | 0.030 | 0.17 |
| *BRCA2* | c.8187G>T (p.Lys2729Asn) | 67 | 60 | 1.03 | (0.72-1.47) | 0.86 | 0.63 |
| *BRCA2* | c.8567A>C (p.Glu2856Ala) | 133 | 172 | 0.89 | (0.70-1.13) | 0.32 | 0.56 |
| *BRCA2* | c.8851G>A (p.Ala2951Thr) | 299 | 340 | 1.04 | (0.88-1.23) | 0.62 | 0.24 |
| *CHEK2* | c.470T>C (p.Ile157Thr) | 520 | 438 | 1.24 | (1.09-1.42) | 0.0013 | 0.0028 |

| Gene | Variant | Case carriers | Control carriers | OR | 95% CI | *P*-value | $P_{ALL}$[a] |
|---|---|---|---|---|---|---|---|
| *CHEK2* | c.538C>T (p.Arg180Cys) | 140 | 124 | 1.44 | (1.12-1.84) | 0.0040 | 0.0016 |
| *PALB2* | c.2816T>G (p.Leu939Trp) | 89 | 100 | 1.03 | (0.77-1.39) | 0.84 | 0.25 |
| *PALB2* | c.2590C>T (p.Pro864Ser) | 103 | 163 | 0.88 | (0.68-1.14) | 0.33 | 0.40 |
| *PALB2* | c.925A>G (p.Ile309Val) | 89 | 63 | 1.33 | (0.95-1.84) | 0.092 | 0.42 |

[a] P-value for association in all training samples

Additional Table 8: P-values for goodness-of-fit tests performed in validation dataset

| | Chi-squared test *P*-value | | |
|---|---|---|---|
| Gene | Logistic regression model[†] | Mixture model with fixed $\alpha^a$ | Mixture model with fixed variant probabilities[b] |
| *ATM* | 0.28 | 0.56 | 0.69 |
| *BRCA1* | 0.74 | 0.43 | 0.40 |
| *BRCA2* | 0.77 | 0.56 | 0.48 |
| *CHEK2* | 0.79 | 0.76 | 0.85 |
| *PALB2* | 0.87 | 0.76 | 0.68 |

*P*-value from chi-squared test comparing expected and observed numbers of cases and controls by risk category[a] or posterior probability interval[b] in the validation data set

**Additional Note**

**Breast Cancer Association Consortium funding and acknowledgements**

FUNDING

A.B Spurdle, M.T. Parsons and C. Fortuno were supported by funding from the Australian National Medical Research Council (IDs 1177524, 1101400, 1161589). The **ABCS** study was supported by the Dutch Cancer Society [grants NKI 2007-3839; 2009 4363]. The **ACP** study is funded by the Breast Cancer Research Trust, UK. KM and AL are supported by the NIHR Manchester Biomedical Research Centre, the Allan Turing Institute under the EPSRC grant EP/N510129/1. The work of the **BBCC** was partly funded by ELAN-Fond of the University Hospital of Erlangen. For **BIGGS**, ES is supported by NIHR Comprehensive Biomedical Research Centre, Guy's & St. Thomas' NHS Foundation Trust in partnership with King's College London, United Kingdom. IT is supported by the Oxford Biomedical Research Centre. The BREast Oncology GAlician Network (**BREOGAN**) is funded by Acción Estratégica de Salud del Instituto de Salud Carlos III FIS PI12/02125/Cofinanciado and FEDER PI17/00918/Cofinanciado FEDER; Acción Estratégica de Salud del Instituto de Salud Carlos III FIS Intrasalud (PI13/01136); Programa Grupos Emergentes, Cancer Genetics Unit, Instituto de Investigacion Biomedica Galicia Sur. Xerencia de Xestion Integrada de Vigo-SERGAS, Instituto de Salud Carlos III, Spain; Grant 10CSA012E, Consellería de Industria Programa Sectorial de Investigación Aplicada, PEME I + D e I + D Suma del Plan Gallego de Investigación, Desarrollo e Innovación Tecnológica de la Consellería de Industria de la Xunta de Galicia, Spain; Grant EC11-192. Fomento de la Investigación Clínica Independiente, Ministerio de Sanidad, Servicios Sociales e Igualdad, Spain; and Grant FEDER-Innterconecta. Ministerio de Economia y Competitividad, Xunta de Galicia, Spain. The **BSUCH** study was supported by the Dietmar-Hopp Foundation, the Helmholtz Society and the German Cancer Research Center (DKFZ). **CCGP** is supported by funding from the University of Crete. The **CECILE** study was supported by Fondation de France, Institut National du Cancer (INCa), Ligue Nationale contre le Cancer, Agence Nationale de Sécurité Sanitaire, de l'Alimentation, de l'Environnement et du Travail (ANSES), Agence Nationale de la Recherche (ANR). The **CGPS** was supported by the Chief Physician Johan Boserup and Lise Boserup Fund, the Danish Medical Research Council, and Herlev and Gentofte Hospital. The **CNIO-BCS** was supported by the Instituto de Salud Carlos III, the Red Temática de Investigación Cooperativa en Cáncer and grants from the Asociación Española Contra el Cáncer and the Fondo de Investigación Sanitario (PI11/00923 and PI12/00070). **COLBCCC** is supported by the German Cancer Research Center (DKFZ), Heidelberg, Germany. Diana Torres was in part supported by a postdoctoral fellowship from the Alexander von Humboldt Foundation. **FHRISK** and **PROCAS** are funded from NIHR grant PGfAR 0707-10031. DGE, AH and WGN are supported by the NIHR Manchester Biomedical Research Centre (IS-BRC-1215-20007). The **GC-HBOC** (German Consortium of Hereditary Breast and Ovarian Cancer) is supported by the German Cancer Aid (grant no 110837, coordinator: Rita K. Schmutzler, Cologne). This work was also funded by the European Regional Development Fund and Free State of Saxony, Germany (LIFE - Leipzig Research Centre for Civilization Diseases, project numbers 713-241202, 713-241202, 14505/2470, 14575/2470). The **GENICA** was funded by the Federal Ministry of Education and Research (BMBF) Germany grants 01KW9975/5, 01KW9976/8, 01KW9977/0 and 01KW0114, the Robert Bosch Foundation, Stuttgart,

Deutsches Krebsforschungszentrum (DKFZ), Heidelberg, the Institute for Prevention and Occupational Medicine of the German Social Accident Insurance, Institute of the Ruhr University Bochum (IPA), Bochum, as well as the Department of Internal Medicine, Evangelische Kliniken Bonn gGmbH, Johanniter Krankenhaus, Bonn, Germany. Generation Scotland (**GENSCOT**) received core support from the Chief Scientist Office of the Scottish Government Health Directorates [CZD/16/6] and the Scottish Funding Council [HR03006]. Genotyping of the GS:SFHS samples was carried out by the Genetics Core Laboratory at the Edinburgh Clinical Research Facility, University of Edinburgh, Scotland and was funded by the Medical Research Council UK and the Wellcome Trust (Wellcome Trust Strategic Award "STratifying Resilience and Depression Longitudinally" (STRADL) Reference 104036/Z/14/Z). Funding for identification of cases and contribution to BCAC funded in part by the Wellcome Trust Seed Award "Temporal trends in incidence and mortality of molecular subtypes of breast cancer to inform public health, policy and prevention" Reference 207800/Z/17/Z. The **GESBC** was supported by the Deutsche Krebshilfe e. V. [70492] and the German Cancer Research Center (DKFZ). The **HABCS** study was supported by the Claudia von Schilling Foundation for Breast Cancer Research, by the Lower Saxonian Cancer Society, and by the Rudolf Bartling Foundation. The **HEBCS** was financially supported by the Helsinki University Hospital Research Fund, the Sigrid Juselius Foundation and The Cancer Foundation Finland. The **HEBON** study is supported by the Dutch Cancer Society grants NKI1998-1854, NKI2004-3088, NKI2007-3756, the Netherlands Organisation of Scientific Research grant NWO 91109024, the Pink Ribbon grants 110005 and 2014-187.WO76, the BBMRI grant NWO 184.021.007/CP46 and the Transcan grant JTC 2012 Cancer 12-054. The **HMBCS** was supported by a grant from the Friends of Hannover Medical School and by the Rudolf Bartling Foundation. The **HUBCS** was supported by a grant from the German Federal Ministry of Research and Education (RUS08/017), B.M. was supported by grant 17-44-020498, 17-29-06014 of the Russian Foundation for Basic Research, D.P. was supported by grant 18-29-09129 of the Russian Foundation for Basic Research, E.K was supported by the program for support the bioresource collections №007-030164/2 and by the megagrant from the Government of Russian Federation No. 075-15-2021-595, and the study was performed as part of the assignment of the Ministry of Science and Higher Education of the Russian Federation (№AAAA-A16-116020350032-1). Financial support for **KARBAC** was provided through the regional agreement on medical training and clinical research (ALF) between Stockholm County Council and Karolinska Institutet, the Swedish Cancer Society, The Gustav V Jubilee foundation and Bert von Kantzows foundation. The **KARMA** study was supported by Märit and Hans Rausings Initiative Against Breast Cancer. The **KBCP** was financially supported by the special Government Funding (VTR) of Kuopio University Hospital grants, Cancer Fund of North Savo, the Finnish Cancer Organizations, and by the strategic funding of the University of Eastern Finland. **kConFab** is supported by a grant from the National Breast Cancer Foundation, and previously by the National Health and Medical Research Council (NHMRC), the Queensland Cancer Fund, the Cancer Councils of New South Wales, Victoria, Tasmania and South Australia, and the Cancer Foundation of Western Australia. Financial support for the AOCS was provided by the United States Army Medical Research and Materiel Command [DAMD17-01-1-0729], Cancer Council Victoria, Queensland Cancer Fund, Cancer Council New South Wales, Cancer Council South Australia, The Cancer Foundation of Western Australia, Cancer Council Tasmania and the National Health and Medical Research Council of Australia (NHMRC; 400413, 400281, 199600). G.C.T. and P.W. are supported by the NHMRC. RB was a Cancer Institute NSW

Clinical Research Fellow. The **KOHBRA** study was partially supported by a grant from the Korea Health Technology R&D Project through the Korea Health Industry Development Institute (KHIDI), and the National R&D Program for Cancer Control, Ministry of Health & Welfare, Republic of Korea (HI16C1127; 1020350; 1420190). The **MARIE** study was supported by the Deutsche Krebshilfe e.V. [70-2892-BR I, 106332, 108253, 108419, 110826, 110828], the Hamburg Cancer Society, the German Cancer Research Center (DKFZ) and the Federal Ministry of Education and Research (BMBF) Germany [01KH0402]. The **MASTOS** study was supported by "Cyprus Research Promotion Foundation" grants 0104/13 and 0104/17, and the Cyprus Institute of Neurology and Genetics. **MBCSG** is supported by grants from the Italian Association for Cancer Research (AIRC). The Melbourne Collaborative Cohort Study (**MCCS**) cohort recruitment was funded by VicHealth and Cancer Council Victoria. The MCCS was further augmented by Australian National Health and Medical Research Council grants 209057, 396414 and 1074383 and by infrastructure provided by Cancer Council Victoria. Cases and their vital status were ascertained through the Victorian Cancer Registry and the Australian Institute of Health and Welfare, including the National Death Index and the Australian Cancer Database. **MYBRCA** is funded by research grants from the Wellcome Trust (v203477/Z/16/Z), the Malaysian Ministry of Higher Education (UM.C/HlR/MOHE/06) and Cancer Research Malaysia. The **NBCS** has received funding from the K.G. Jebsen Centre for Breast Cancer Research; the Research Council of Norway grant 193387/V50 (to A-L Børresen-Dale and V.N. Kristensen) and grant 193387/H10 (to A-L Børresen-Dale and V.N. Kristensen), South Eastern Norway Health Authority (grant 39346 to A-L Børresen-Dale) and the Norwegian Cancer Society (to A-L Børresen-Dale and V.N. Kristensen). The Ontario Familial Breast Cancer Registry (**OFBCR**) was supported by grant U01CA164920 from the USA National Cancer Institute of the National Institutes of Health. The content of this manuscript does not necessarily reflect the views or policies of the National Cancer Institute or any of the collaborating centers in the Breast Cancer Family Registry (BCFR), nor does mention of trade names, commercial products, or organizations imply endorsement by the USA Government or the BCFR. The **ORIGO** study was supported by the Dutch Cancer Society (RUL 1997-1505) and the Biobanking and Biomolecular Resources Research Infrastructure (BBMRI-NL CP16). The **PBCS** was funded by Intramural Research Funds of the National Cancer Institute, Department of Health and Human Services, USA. Genotyping for PLCO was supported by the Intramural Research Program of the National Institutes of Health, NCI, Division of Cancer Epidemiology and Genetics. The **PLCO** is supported by the Intramural Research Program of the Division of Cancer Epidemiology and Genetics and supported by contracts from the Division of Cancer Prevention, National Cancer Institute, National Institutes of Health. The **RBCS** was funded by the Dutch Cancer Society (DDHK 2004-3124, DDHK 2009-4318). The **SASBAC** study was supported by funding from the Agency for Science, Technology and Research of Singapore (A*STAR), the US National Institute of Health (NIH) and the Susan G. Komen Breast Cancer Foundation. **SEARCH** is funded by Cancer Research UK [C490/A10124, C490/A16561] and supported by the UK National Institute for Health Research Biomedical Research Centre at the University of Cambridge. The University of Cambridge has received salary support for PDPP from the NHS in the East of England through the Clinical Academic Reserve. **SGBCC** is funded by the National Research Foundation Singapore (NRF-NRFF2017-02), NUS start-up Grant, National University Cancer Institute Singapore (NCIS) Centre Grant, Breast Cancer Prevention Programme, Asian Breast Cancer Research Fund and the NMRC Clinician Scientist Award (SI Category). Population-based controls were from the Multi-Ethnic Cohort (MEC) funded by grants from the Ministry of Health, Singapore, National University of Singapore and National University Health System, Singapore. **SKKDKFZS** is supported by the DKFZ. The **SZBCS** was supported by Grant PBZ_KBN_122/P05/2004 and the program of the Minister of Science and Higher Education under the name "Regional Initiative of Excellence" in 2019-2022 project number 002/RID/2018/19 amount of financing 12 000 000 PLN. **UBCS** was supported by funding from National Cancer Institute (NCI) grant R01 CA163353 (to N.J. Camp) and the Women’s Cancer Center at the Huntsman Cancer Institute (HCI). Data collection for UBCS was supported by the Utah Population Database (UPDB) and Utah Cancer Registry (UCR). The UPDB is supported by HCI (including the Huntsman Cancer Foundation), University of Utah program in Personalized Health and Center for Clinical and Translational Science, and NCI grant P30 CA2014. The UCR is funded by the NCI's SEER Program, Contract No. HHSN261201800016I, the US Center for Disease Control and Prevention's National Program of Cancer Registries, Cooperative Agreement No. NU58DP0063200, the University of Utah and Huntsman Cancer Foundation.

ACKNOWLEDGEMENTS

We thank all the individuals who took part in these studies and all the researchers, clinicians, technicians and administrative staff who have enabled this work to be carried out. **ABCS** thanks the Blood bank Sanquin, The Netherlands. The **ACP** study wishes to thank the participants in the Thai Breast Cancer study. Special thanks also go to the Thai Ministry of Public Health (MOPH), doctors and nurses who helped with the data collection process. Finally, the study would like to thank Dr Prat Boonyawongviroj, the former Permanent Secretary of MOPH and Dr Pornthep Siriwanarungsan, the former Department Director-General of Disease Control who have supported the study throughout. **BBCS** thanks Eileen Williams, Elaine Ryder-Mills, Kara Sargus. **BIGGS** thanks Niall McInerney, Gabrielle Colleran, Andrew Rowan, Angela Jones. The **BREOGAN** study would not have been possible without the contributions of the following: Manuela Gago-Dominguez, Jose Esteban Castelao, Angel Carracedo, Victor Muñoz Garzón, Alejandro Novo Domínguez, Maria Elena Martinez, Sara Miranda Ponte, Carmen Redondo Marey, Maite Peña Fernández, Manuel Enguix Castelo, Maria Torres, Manuel Calaza (BREOGAN), José Antúnez, Máximo Fraga and the staff of the Department of Pathology and Biobank of the University Hospital Complex of Santiago-CHUS, Instituto de Investigación Sanitaria de Santiago, IDIS, Xerencia de Xestion Integrada de Santiago-SERGAS; Joaquín González-Carreró and the staff of the Department of Pathology and Biobank of University Hospital Complex of Vigo, Instituto de Investigacion Biomedica Galicia Sur, SERGAS, Vigo, Spain. The **BSUCH** study acknowledges the Principal Investigator, Barbara Burwinkel, and, thanks Peter Bugert, Medical Faculty Mannheim. **CCGP** thanks Styliani Apostolaki, Anna Margiolaki, Georgios Nintos, Maria Perraki, Georgia Saloustrou, Georgia Sevastaki, Konstantinos Pompodakis. **CGPS** thanks staff and participants of the Copenhagen General Population Study. For the excellent technical assistance: Dorthe Uldall Andersen, Maria Birna Arnadottir, Anne Bank, Dorthe Kjeldgård Hansen. The Danish Cancer Biobank is acknowledged for providing infrastructure for the collection of blood samples for the cases. **CNIO-BCS** thanks Guillermo Pita, Charo Alonso, Nuria Álvarez, Pilar Zamora, Primitiva Menendez, the Human Genotyping-CEGEN Unit (CNIO). **COLBCCC** thanks all patients, the physicians Justo G. Olaya, Mauricio Tawil, Lilian Torregrosa, Elias Quintero, Sebastian Quintero, Claudia Ramírez, José J. Caicedo, and Jose F. Robledo, and the technician Michael Gilbert for their contributions and commitment to this study. **FHRISK** and **PROCAS** thank NIHR for funding. **GC-HBOC** thanks Stefanie Engert, Heide Hellebrand, Sandra Kröber and LIFE - Leipzig Research Centre for Civilization Diseases (Markus Loeffler, Joachim Thiery, Matthias Nüchter, Ronny Baber). The **GENICA** Network: Dr. Margarete Fischer-Bosch-Institute of Clinical Pharmacology, Stuttgart, and University of Tübingen, Germany [Hiltrud Brauch, Wing-Yee Lo, Reiner Hoppe], German Cancer Consortium (DKTK) and German Cancer Research Center (DKFZ), Partner Site Tübingen, 72074 Tübingen, Germany [Hiltrud Brauch], gefördert durch die Deutsche Forschungsgemeinschaft (DFG) im Rahmen der Exzellenzstrategie des Bundes und der Länder - EXC 2180 - 390900677 [Hiltrud Brauch], Department of Internal Medicine, Evangelische Kliniken Bonn gGmbH, Johanniter Krankenhaus, Bonn, Germany [YDK, Christian Baisch], Institute of Pathology, University of Bonn, Germany [Hans-Peter Fischer], Molecular Genetics of Breast Cancer, Deutsches Krebsforschungszentrum (DKFZ), Heidelberg, Germany [Ute Hamann], Institute for Prevention and Occupational Medicine of the German Social Accident Insurance, Institute of the Ruhr University Bochum (IPA), Bochum, Germany [Thomas Brüning, Beate Pesch, Sylvia Rabstein, Anne Lotz]; and Institute of Occupational Medicine and Maritime Medicine, University Medical Center Hamburg-Eppendorf, Germany [Volker Harth]. **HABCS** thanks Michael Bremer. **HEBCS** thanks Carl Blomqvist, Taru A. Muranen, Kristiina Aittomäki, Outi Malkavaara. **HEBON** Investigators are J. Margriet Collée, Frans B. L. Hogervorst, Maartje J. Hooning, Carolien M. Kets, Peter Devilee, Christi J. van Asperen, Matti A. Rookus, Marjanka K. Schmidt, Cora M. Aalfs, Muriel A. Adank, Margreet G. E. M. Ausems, Marinus J. Blok, Encarna B. Gómez Garcia, Bernadette A. M. Heemskerk-Gerritsen, Antoinette Hollestelle, Agnes Jager, Linetta B. Koppert, Marco Koudijs, Mieke Kriege, Hanne E. J. Meijers-Heijboer, Arjen R. Mensenkamp, Thea M. Mooij, Jan C. Oosterwijk, Ans M. W. van den Ouweland, Frederieke H. van der Baan, Annemieke H. van der Hout, Lizet E. van der Kolk, Rob B. van der Luijt, Carolien H. M. van Deurzen, Helena C. van Doorn, Klaartje van Engelen, Liselotte P. van Hest, Theo A. M. van Os, Senno Verhoef, Maartje J. Vogel & Juul T. Wijnen. **HMBCS** thanks Peter Hillemanns, Hans Christiansen and Johann H. Karstens. **HUBCS** thanks Darya Prokofyeva and Shamil Gantsev. **KARMA** and **SASBAC** thank the Swedish Medical Research Counsel. **KBCP** thanks Eija Myöhänen. **kConFab/AOCS** wish to thank Heather Thorne, Eveline Niedermayr, all the kConFab research nurses and staff, the heads and staff of the Family Cancer Clinics, and the Clinical Follow Up Study (which has received funding from the NHMRC, the National Breast Cancer Foundation, Cancer Australia, and the National Institute of Health (USA)) for their contributions to this resource, and the many families who contribute to kConFab. We thank all investigators of the **KOHBRA** (Korean Hereditary Breast Cancer) Study. **MARIE** thanks Petra Seibold, Nadia Obi, Sabine Behrens, Ursula Eilber and Muhabbet Celik. **MASTOS** thanks all the study participants and express appreciation to the doctors: Yiola Marcou, Eleni Kakouri, Panayiotis Papadopoulos, Simon Malas and Maria Daniel, as well as to all the nurses and volunteers who provided valuable help towards the recruitment of the study participants. **MBCSG** (Milan Breast Cancer Study Group): Manoukian Siranoush, Bernard Peissel, Jacopo Azzollini, Daniela Zaffaroni, Bernardo Bonanni, Irene Feroce, Mariarosaria Calvello, Aliana Guerrieri Gonzaga, Monica Marabelli, Davide Bondavalli and the personnel of the Cogentech Cancer Genetic Test Laboratory. The **MCCS** was made possible by the contribution of many people, including the original investigators, the teams that recruited the participants and continue working on follow-up, and the many thousands of Melbourne residents who continue to participate in the study. We thank the coordinators, the research staff and especially the **MYBRCA** thanks study participants and research staff (particularly Patsy Ng, Nurhidayu Hassan, Yoon Sook-Yee,

Daphne Lee, Lee Sheau Yee, Phuah Sze Yee and Norhashimah Hassan) for their contributions and commitment to this study. The **OFBCR** thanks Teresa Selander, Nayana Weerasooriya and Steve Gallinger. **ORIGO** thanks E. Krol-Warmerdam, and J. Blom for patient accrual, administering questionnaires, and managing clinical information. The LUMC survival data were retrieved from the Leiden hospital-based cancer registry system (ONCDOC) with the help of Dr. J. Molenaar. **PBCS** thanks Louise Brinton, Mark Sherman, Neonila Szeszenia-Dabrowska, Beata Peplonska, Witold Zatonski, Pei Chao, Michael Stagner. We thank the **SEARCH** and **EPIC** teams. **SGBCC** thanks the participants and all research coordinators for their excellent help with recruitment, data and sample collection. **SKKDKFZ**S thanks all study participants, clinicians, family doctors, researchers and technicians for their contributions and commitment to this study. **SZBCS** thanks Ewa Putresza. **UBCS** thanks all study participants, the ascertainment, laboratory and research informatics teams at Huntsman Cancer Institute and Intermountain Healthcare, and Justin Williams, Brandt Jones, Melissa Cessna, Stacey Knight and Kerry Rowe for their important contributions to this study.
